# Effect of stem cell infusion timing on acute graft-versus-host disease: a randomized clinical trial

**DOI:** 10.64898/2026.05.03.26352313

**Authors:** Yue Wu, Jiayu Huang, Yang Yang, Weijie Cao, Yanmin Zhao, Yujun Dong, Weiwei Wu, Xiaoxia Hu, Yang Cao, Ran Zhang, Congxiao Zhang, Bingjie Wang, Kaidi Song, Guangyu Sun, Wen Yao, Qing Cheng, Jian Wang, Meijuan Tu, Yiwen Hou, Cheng Zhan, Xiaoyu Zhu

## Abstract

**BACKGROUND:** Our previous retrospective study suggested that peripheral-blood stem-cell infusion before 2:00 p.m. was associated with a lower incidence of acute graft-versus-host disease (aGVHD) than later infusion, but retrospective analyses are susceptible to confounding. Whether infusion timing affects aGVHD has not been tested in a randomized trial.

**METHODS:** In this multicenter, open-label trial, we randomly assigned patients 12 to 60 years of age with hematologic malignancies undergoing a first allogeneic peripheral-blood stem-cell transplantation to stem-cell infusion at 12:00 pm (±30 minutes) or 6:00 pm (±30 minutes). The primary end point was grade II to IV aGVHD within 100 days after transplantation.

**RESULTS:** A total of 198 patients underwent randomization (99 in each group). Grade II to IV acute GVHD occurred in 11.1% of patients in the 12:00 pm group and 23.2% in the 6:00 pm group (subdistribution hazard ratio for 6:00 pm vs. 12:00 pm, 2.18; 95% CI, 1.06 to 4.48; P = 0.029). The corresponding incidences of grade III to IV acute GVHD were 2.0% and 12.2% (subdistribution hazard ratio, 6.25; 95% CI, 1.39 to 28.15; P = 0.006). There were no significant differences in hematopoietic recovery, TRM, or relapse between groups.

**CONCLUSIONS:** Among patients undergoing allogeneic peripheral-blood stem-cell transplantation, infusion at 12:00 pm resulted in a lower incidence of grade II to IV acute GVHD than infusion at 6:00 pm (ClinicalTrials.gov number, NCT06294678.)

## Introduction

Allogeneic hematopoietic stem cell transplantation (allo-HSCT) has expanded rapidly over the past two decades and remains an established curative therapy for patients with malignant hematologic diseases.^1^ As transplant numbers rise, the prevention and management of transplantation-related complications have become critical determinants of long-term outcomes.^2,3^ Acute graft-versus-host disease (aGVHD) is a major post-transplantation complication that drastically reduces quality of life and is a leading cause of transplantation-related mortality (TRM). Despite advances in conditioning regimens and GVHD prophylaxis, the incidence of grade II-IV aGVHD remains high, ranging from approximately 14% to 47%, depending on donor source and transplant setting.^4,5^ Importantly, severe aGVHD (grade III-IV) is a key driver of poor survival, with a reported 3-year overall survival (OS) of approximately 29% in affected patients.^4^ Consequently, aGVHD continues to limit the long-term benefit of allo-HSCT.

Chronotherapy has gained increasing attention across a broad range of clinical settings.^6–8^ Recent retrospective analyses and preclinical studies have demonstrated that the circadian timing of therapeutic intervention significantly affects treatment outcomes^9,10^, including anti-PD-1 immunotherapy for melanoma or lung cancer.^11–13^ Notably, stem cell infusion time-of-day has been linked to the risk of aGVHD and survival after transplantation. In our retrospective cohorts of allogeneic peripheral blood stem cell transplantation (allo-PBSCT), infusion earlier in the day (< 2:00 pm) was associated with significantly lower incidences of grade II-IV and grade III-IV aGVHD, as well as superior GVHD-free, relapse-free survival (GRFS), compared with infusion later in the day (≥ 2:00 pm).^14^ Among recipients of cryopreserved unrelated cord blood transplantation, graft infusion before 9:40 am was associated with a reduced risk of severe aGVHD.^15^ Another independent retrospective study further confirmed that patients receiving allo-PBSCT before 11:00 am had significantly better 2-year overall survival (OS) and lower TRM than those infused later in the day.^16^

Although these retrospective studies consistently suggest that optimizing the timing of stem cell infusion reduces aGVHD, improves survival and enhances transplant outcomes, findings from retrospective investigations are inherently susceptible to confounding bias. For instance, two recent retrospective studies exploring the impact of infusion timing on the efficacy of CAR-T therapy in large B-cell lymphoma reported contradictory results.^17,18^ Therefore, prospective, randomized controlled trials are urgently needed to validate whether circadian timing of graft infusion truly improves clinical outcomes after allo-HSCT.

Here, we report the results of a randomized clinical trial (RCT), which evaluating the effect of stem cell infusion time-of-day on the incidence and severity of aGVHD in patients undergoing allo-PBSCT.

## Methods

### Trial Design

This investigator-initiated, multicenter, prospective, randomized, open-label, phase 3 clinical trial was conducted at 6 centers in China (see the Supplementary Appendix). It evaluated the effect of stem cell infusion time-of-day on the incidence and severity of aGVHD in patients undergoing their first allo-PBSCT. The corresponding authors conceived the trial. The first author designed the protocol and prepared the initial draft, with input from the other authors. The study protocol was approved by the institutional review board at the coordinating center and by the ethics committees at all participating sites (REC reference: 2024/KY/074) (Data S1). The trial was conducted in accordance with the Declaration of Helsinki, the International Council for Harmonization Guideline for Good Clinical Practice and the CONSORT guideline. Written informed consent was obtained from all participants or their legal representatives.

### Patients and Randomization

Eligible patients were aged 12 to 60 years with a confirmed diagnosis of malignant hematologic disease, as defined by the World Health Organization Classification of Haematolymphoid Tumours (WHO-HAEM5), and were scheduled to undergo first-time allo-PBSCT. Transplantation donors included matched sibling donors, haploidentical related donors, and unrelated donors. Key exclusion criteria included prior HSCT and participation in other clinical trials that could affect aGVHD outcomes within 3 months. Full eligibility criteria are provided in the trial protocol (Data S1-Protocol).

Eligible patients were randomly assigned in a 1:1 ratio to receive stem cell infusion either at an earlier time-of-day (12:00 pm ± 0.5 hour) or a later time-of-day (6:00 pm ± 0.5 hour). Randomization was performed centrally using a computer-generated sequence with randomly varying permuted block sizes of 4, 6, and 8, stratified according to the study center. Allocation was implemented through an electronic data capture (EDC) system, ensuring concealment of group assignments until assignment.

### Trial Procedures

Conditioning regimens were not standardized and included both myeloablative conditioning (MAC) and reduced-intensity conditioning (RIC) regimens administered according to institutional practice. Common regimens consisted of busulfan-based regimens, which were combined with cyclophosphamide or melphalan, fludarabine or cytarabine, and total body irradiation or total marrow irradiation-based regimens with cyclophosphamide and additional cytarabine. The target stem cell doses for collection and infusion were approximately 5×10 total nucleated cells (TNCs) per kilogram of recipient body weight and at least 2×10 CD34^+^ cells per kilogram.

According to randomized assignment, stem cell infusion was protocol-specified to start at either 12:00 pm (± 0.5 hour) or 6:00 pm (± 0.5 hour). For patients in whom the total planned cell dose could not be delivered in a single session, additional infusion was permitted on the following day within the same assigned time window. Transplantation day 0 was defined as the date of the first infusion. Patients whose infusion start time fell outside the predefined window were excluded from the per-protocol analysis.

Standard prophylaxis for aGVHD consisted of calcineurin inhibitors (CNIs, cyclosporine or tacrolimus) in combination with mycophenolate mofetil (MMF), with additional use of rabbit antithymocyte globulin (rATG) or anti-T-lymphocyte globulin (ATLG), post-transplant cyclophosphamide (PT-Cy), and/or short-course methotrexate (MTX) according to donor type and institutional practice. These regimens were applied according to predefined institutional protocols across study centers, with details provided in the Supplementary Appendix. Supportive care, including antimicrobial prophylaxis and growth factor support, was provided according to institutional standards at each participating center.

### End Points

The primary end point was the cumulative incidence of grade II to IV aGVHD within 100 days after transplantation, graded according to the Mount Sinai Acute GVHD International Consortium (MAGIC) criteria.^19^ For each suspected aGVHD event, the diagnosis and grade were independently reviewed and confirmed by at least 2 transplant physicians at the participating center. Relevant objective clinical data, including organ involvement, laboratory findings, treatment records, and other source documentation, were entered into the EDC system for verification. The secondary end points included the cumulative incidences of grade III to IV aGVHD, neutrophil engraftment, platelet recovery, TRM, chronic GVHD (cGVHD), graded according to the 2014 National Institutes of Health criteria^20^, and probabilities of GRFS, disease-free survival (DFS) and OS. Comprehensive definitions for all time-to-event end points are detailed in protocol (Data S1).

### Statistical Analysis

The sample size was calculated with G*Power on the basis of a preliminary analysis of 134 matched sibling donor transplant recipients from a previously published 204-patient single-center cohort,^14^ in whom the day-100 incidence of grade II to IV aGVHD was 18.0% with stem-cell infusion before 2:00 pm and 41.0% with infusion at or after 2:00 pm. A total of 178 patients were required to provide 90% power at a two-sided significance level of 0.05 to detect this difference. Allowing for 10% attrition, the target enrollment was 198 patients.

Analyses of the primary and secondary end points were performed in the full analysis set, defined as all randomized patients who underwent transplantation, according to the intention-to-treat principle. Analyses were also conducted in the per-protocol set. aGVHD was assessed from transplantation day 0 through day 100. Clinical outcomes, including GVHD events and survival, were recorded in a standardized EDC system and verified against source medical records. The primary analysis was unadjusted. Prespecified multivariable analyses adjusted for clinically relevant baseline and transplantation-related characteristics. Statistical analyses were conducted according to the prespecified statistical analysis plan; additional details are provided in the Supplementary Appendix.

## Results

### Patients

Between March 18, 2024, and June 11, 2025, a total of 235 patients were screened at 6 centers in China, of whom 37 were excluded, primarily because of nonmalignant hematologic disease (n = 26) or failure to meet age criteria (n = 11). A total of 198 patients underwent randomization, and all received transplantation (Fig. 1). Among all patients, acute myeloid leukemia was the most common diagnosis (50.0%), followed by acute lymphoblastic leukemia (28.8%) and myelodysplastic syndromes/myeloproliferative neoplasms (15.2%); 12 patients had other malignant hematologic diseases. The median age of the cohort was 38 years (range, 13-60), and 119 patients (60.1%) were male. The median TNC dose was 9.30 ×10 /kg (range, 1.32-63.98), and the median infused CD34^+^ cell dose was 6.5 ×10 /kg (range, 2.19-43.77). 127 of 198 (64.1%) patients received MAC, and 71 patients (35.9%) received an RIC regimen. Baseline transplantation characteristics were generally comparable between the two groups (Table 1). Detailed information on the conditioning regimen is shown in Table S1. A total of 15 patients received stem cell infusion on 2 consecutive days because the target cell dose could not be delivered in a single session. All patients received CNIs combined with MMF as the basic aGVHD prophylaxis regimen; 4 patients received tacrolimus, and the remainder received cyclosporine. The database was locked on Jun 30, 2026, for the present analysis. The median follow-up among surviving patients was 655 days (range, 403 to 834 days).

**Figure 1.**
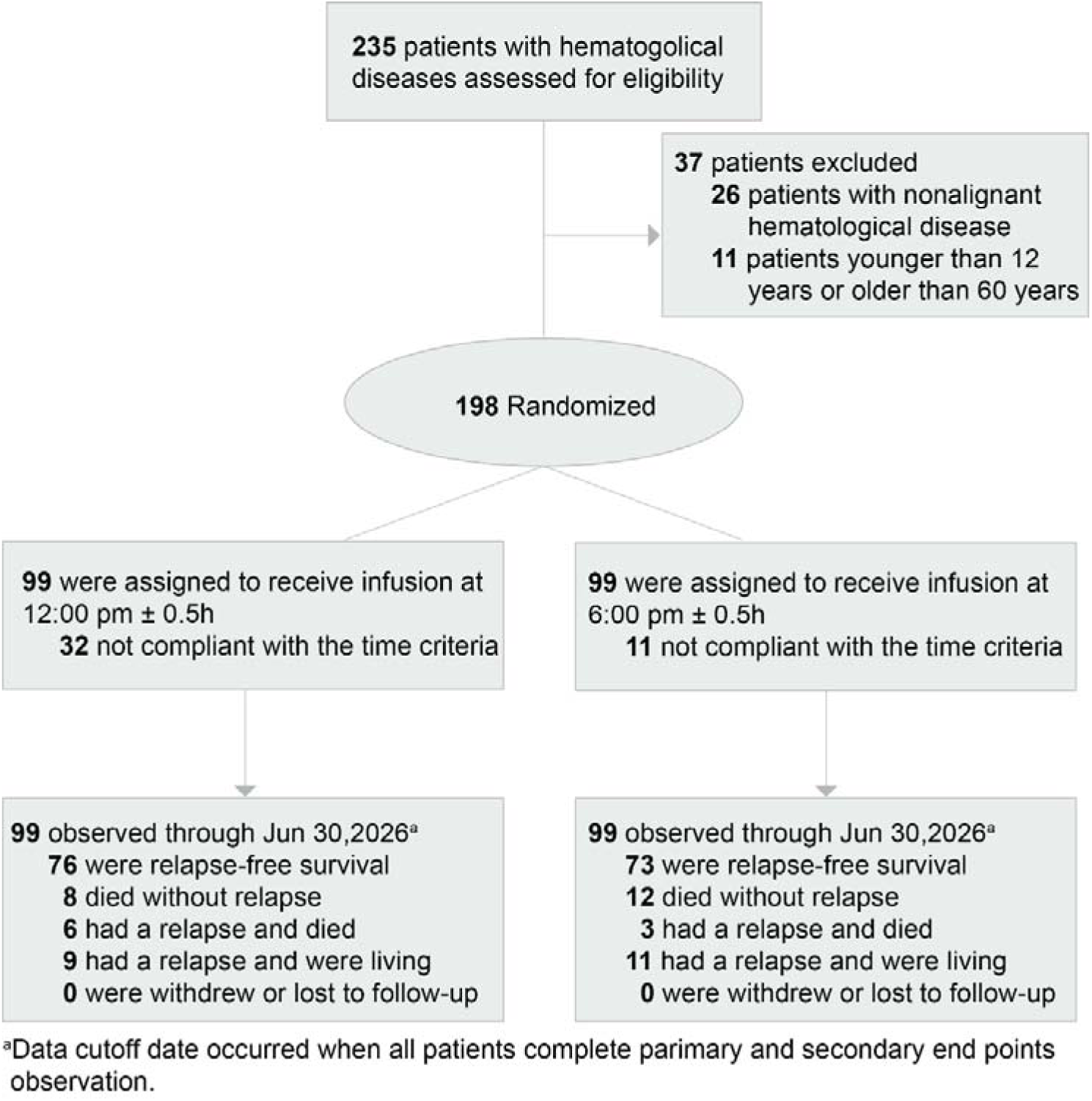
Enrollment, Randomization, Treatment and Follow-up.

**Table 1.**
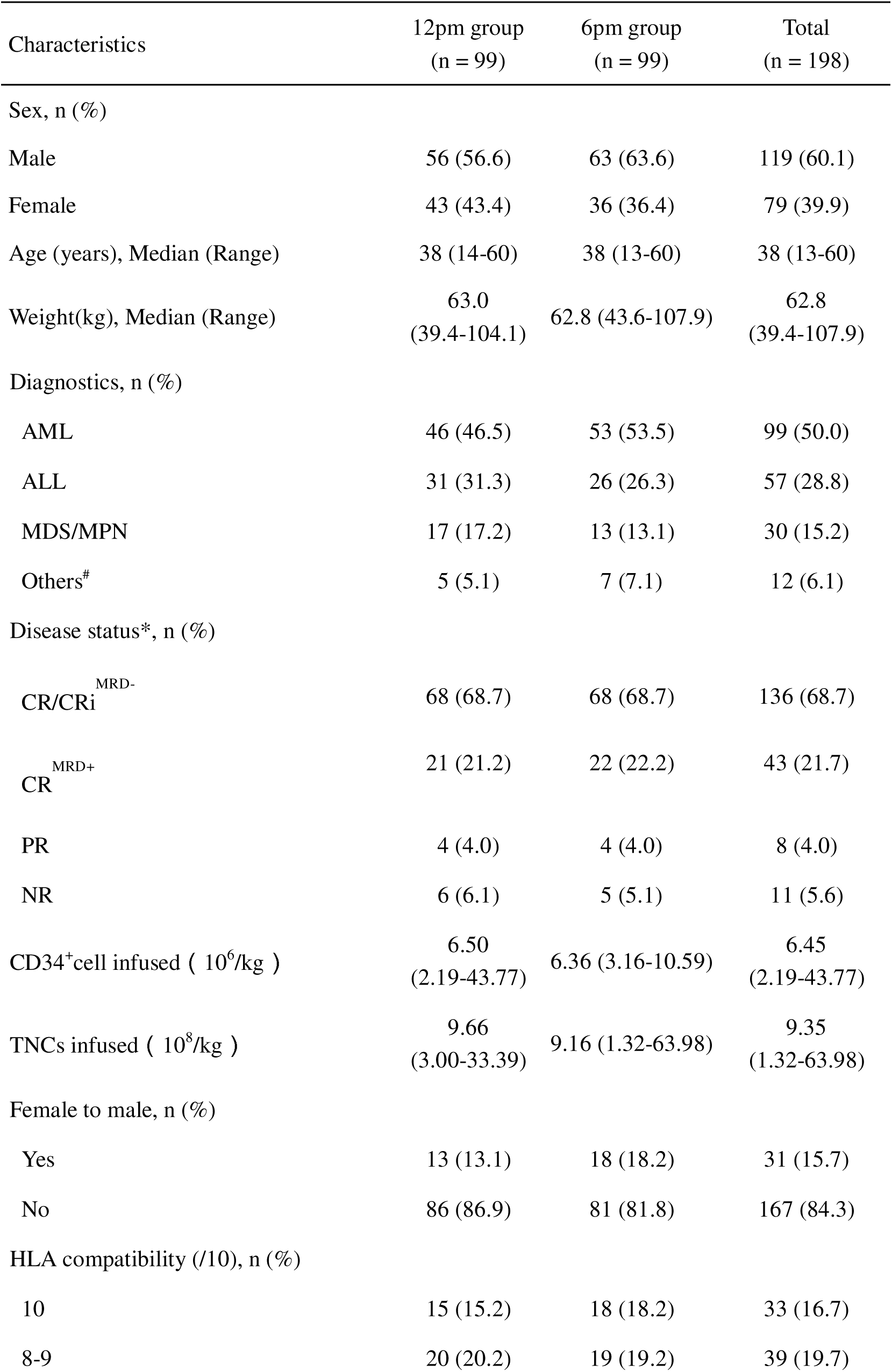
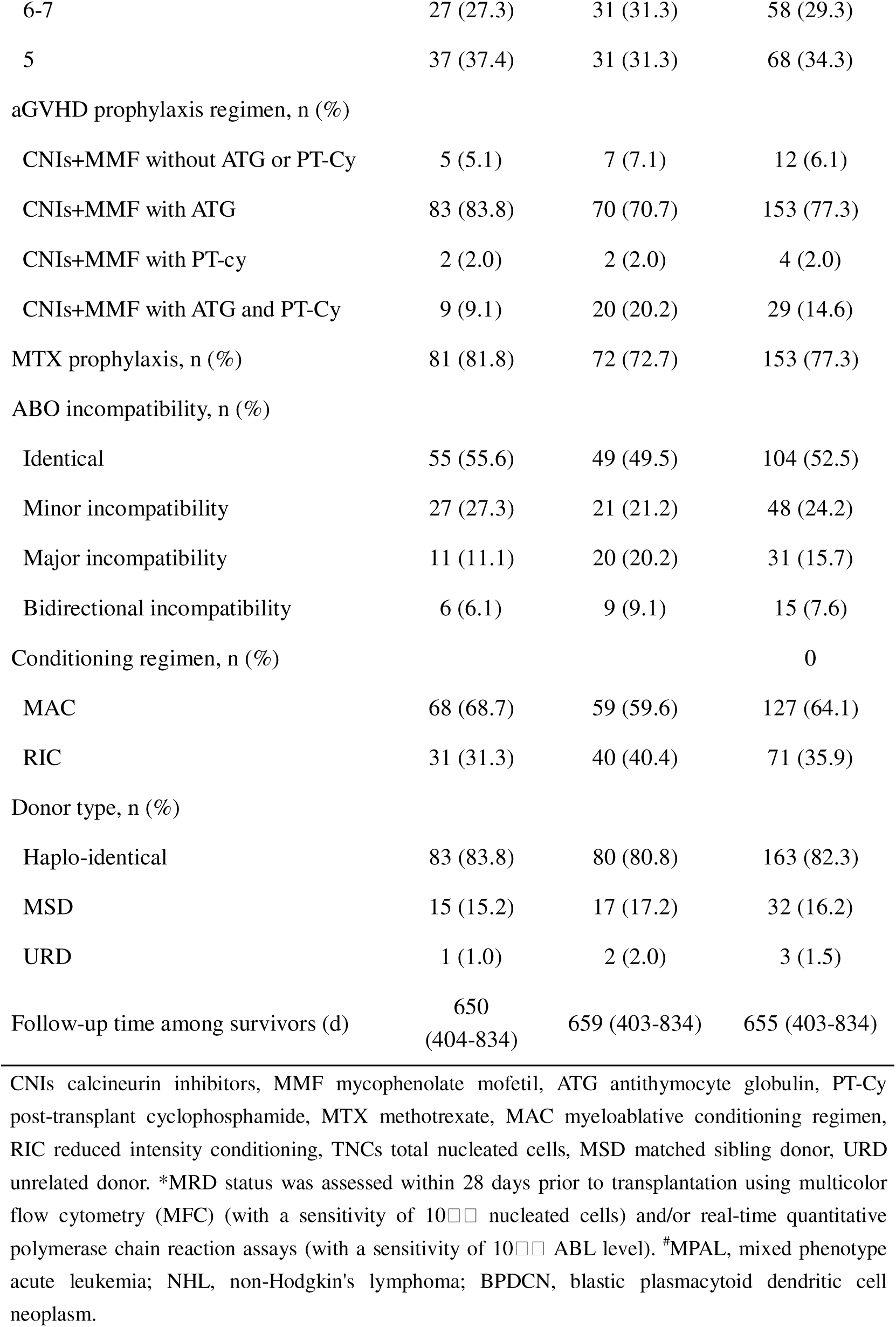
Baseline characteristics of the patients in the intention-to-treat population.

| Characteristics | 12pm group<br>(n = 99) | 6pm group<br>(n = 99) | Total<br>(n = 198) |
| --- | --- | --- | --- |
| Sex, n (%) |  |  |  |
| Male | 56 (56.6) | 63 (63.6) | 119 (60.1) |
| Female | 43 (43.4) | 36 (36.4) | 79 (39.9) |
| Age (years), Median (Range) | 38 (14-60) | 38 (13-60) | 38 (13-60) |
| Weight(kg), Median (Range) | 63.0<br>(39.4-104.1) | 62.8 (43.6-107.9) | 62.8<br>(39.4-107.9) |
| Diagnostics, n (%) |  |  |  |
| AML | 46 (46.5) | 53 (53.5) | 99 (50.0) |
| ALL | 31 (31.3) | 26 (26.3) | 57 (28.8) |
| MDS/MPN | 17 (17.2) | 13 (13.1) | 30 (15.2) |
| Others<sup>#</sup> | 5 (5.1) | 7 (7.1) | 12 (6.1) |
| Disease status*, n (%) |  |  |  |
| CR/CRi<sup>MRD-</sup> | 68 (68.7) | 68 (68.7) | 136 (68.7) |
| CR<sup>MRD+</sup> | 21 (21.2) | 22 (22.2) | 43 (21.7) |
| PR | 4 (4.0) | 4 (4.0) | 8 (4.0) |
| NR | 6 (6.1) | 5 (5.1) | 11 (5.6) |
| CD34<sup>+</sup> cell infused ( 10<sup>6</sup>/kg ) | 6.50<br>(2.19-43.77) | 6.36 (3.16-10.59) | 6.45<br>(2.19-43.77) |
| TNCs infused ( 10<sup>8</sup>/kg ) | 9.66<br>(3.00-33.39) | 9.16 (1.32-63.98) | 9.35<br>(1.32-63.98) |
| Female to male, n (%) |  |  |  |
| Yes | 13 (13.1) | 18 (18.2) | 31 (15.7) |
| No | 86 (86.9) | 81 (81.8) | 167 (84.3) |
| HLA compatibility (/10), n (%) |  |  |  |
| 10 | 15 (15.2) | 18 (18.2) | 33 (16.7) |
| 8-9 | 20 (20.2) | 19 (19.2) | 39 (19.7) |
| 6-7 | 27 (27.3) | 31 (31.3) | 58 (29.3) |
| 5 | 37 (37.4) | 31 (31.3) | 68 (34.3) |
| aGVHD prophylaxis regimen, n (%) |  |  |  |
| CNIs+MMF without ATG or PT-Cy | 5 (5.1) | 7 (7.1) | 12 (6.1) |
| CNIs+MMF with ATG | 83 (83.8) | 70 (70.7) | 153 (77.3) |
| CNIs+MMF with PT-cy | 2 (2.0) | 2 (2.0) | 4 (2.0) |
| CNIs+MMF with ATG and PT-Cy | 9 (9.1) | 20 (20.2) | 29 (14.6) |
| MTX prophylaxis, n (%) | 81 (81.8) | 72 (72.7) | 153 (77.3) |
| ABO incompatibility, n (%) |  |  |  |
| Identical | 55 (55.6) | 49 (49.5) | 104 (52.5) |
| Minor incompatibility | 27 (27.3) | 21 (21.2) | 48 (24.2) |
| Major incompatibility | 11 (11.1) | 20 (20.2) | 31 (15.7) |
| Bidirectional incompatibility | 6 (6.1) | 9 (9.1) | 15 (7.6) |
| Conditioning regimen, n (%) |  |  | 0 |
| MAC | 68 (68.7) | 59 (59.6) | 127 (64.1) |
| RIC | 31 (31.3) | 40 (40.4) | 71 (35.9) |
| Donor type, n (%) |  |  |  |
| Haplo-identical | 83 (83.8) | 80 (80.8) | 163 (82.3) |
| MSD | 15 (15.2) | 17 (17.2) | 32 (16.2) |
| URD | 1 (1.0) | 2 (2.0) | 3 (1.5) |
| Follow-up time among survivors (d) | 650<br>(404-834) | 659 (403-834) | 655 (403-834) |

CNIs calcineurin inhibitors, MMF mycophenolate mofetil, ATG antithymocyte globulin, PT-Cy post-transplant cyclophosphamide, MTX methotrexate, MAC myeloablative conditioning regimen, RIC reduced intensity conditioning, TNCs total nucleated cells, MSD matched sibling donor, URD unrelated donor. \*MRD status was assessed within 28 days prior to transplantation using multicolor flow cytometry (MFC) (with a sensitivity of $10^{-4}$ nucleated cells) and/or real-time quantitative polymerase chain reaction assays (with a sensitivity of $10^{-4}$ ABL level). #MPAL, mixed phenotype acute leukemia; NHL, non-Hodgkin's lymphoma; BPDCN, blastic plasmacytoid dendritic cell neoplasm.

Among randomized patients, the actual infusion start times ranged from 10:32 am to 8:12 pm in the 12:00 pm group and from 2:48 pm to 7:50 pm in the 6:00 pm group (Fig. S1). According to the protocol, 155 patients (78.3%) initiated stem cell infusion within the prespecified time window and were included in the per-protocol set, including 67 patients in the 12:00 pm group and 88 in the 6:00 pm group. Patients were excluded from the per-protocol analysis solely because stem cell infusion was initiated outside the predefined time window. Baseline characteristics were not significantly different between the 12:00 pm and 6:00 pm groups in the per-protocol population (Table S2). Baseline characteristics were also comparable between patients who adhered to the assigned infusion time window and those who did not within each randomized group (Table S3).

Among randomized patients, protocol deviations from the prespecified infusion window occurred more frequently in the 12:00 pm group than in the 6:00 pm group. These deviations were mainly due to logistical delays in stem cell collection and processing rather than patient-related clinical factors. Although 32 patients in the 12:00 pm group initiated infusion outside the predefined 11:30 am to 12:30 pm window, only 14 initiated infusion after 2:00 pm (Fig. S1), indicating that most deviations still occurred within the earlier part of the day.

### Primary End Point

We evaluated the effect of infusion timing in both the intention to treat and per protocol populations, and further performed sensitivity analyses using alternative time cutoffs. In the intention-to-treat population, the cumulative incidence of grade II-IV aGVHD was significantly lower in the 12:00 pm group than in the 6:00 pm group (11.1% [95% CI, 5.9%-18.2%] vs 23.2% [95% CI, 15.4%-32.0%], *P* = 0.029; hazard ratio, 2.18 [95% CI, 1.06-4.48]; Fig. 2A). Grade III aGVHD occurred in 2 patients in each group, whereas grade IV aGVHD occurred only in the 6:00 pm group (10 patients). The multivariable analysis in the intention-to-treat population was performed. The association between infusion timing and aGVHD remained significant after adjustment (12 pm vs. 6 pm, hazard ratio, 2.39 [95% CI, 1.13-5.04]; *P* = 0.023) (Table 2). In the per-protocol set, the cumulative incidence of grade II-IV aGVHD was also significantly lower in the 12:00 pm group (9.0% [95% CI, 3.6%-17.3%] vs 26.1% [95% CI, 17.4%-35.7%], *P* = 0.008; hazard ratio, 3.16 [95% CI, 1.28-7.78]; Fig. S2A). The multivariable analysis showed that later infusion time (6:00 pm group) was independently associated with the occurrence of grade II-IV aGVHD in the per-protocol set (Table 2). In sensitivity analyses using alternative time cutoffs from 1:00 pm to 5:00 pm, later infusion remained associated with a higher risk of aGVHD (Table S4). Organ involvement patterns differed between the two groups, with a higher frequency of gastrointestinal involvement in the 6:00 pm group (Table S5).

**Figure 2.**
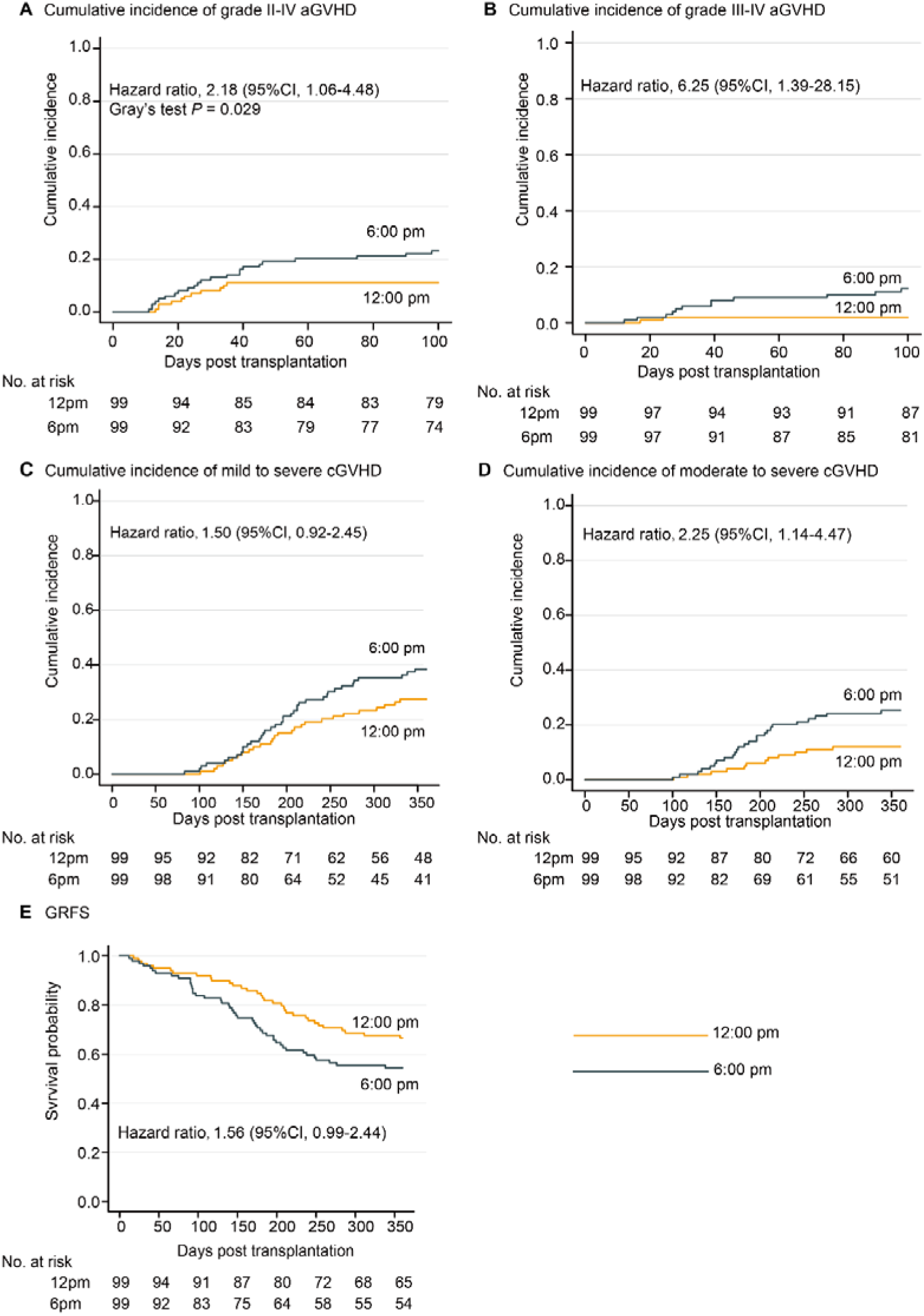
Effects of stem cell infusion timing on GVHD and survival in the intention-to-treat population. (**A** and **B**) Cumulative incidences (*CI*) of grade II-IV aGVHD (**A**) and grade III-IV aGVHD (**B**) in the 12 pm (n = 99) and 6 pm (n = 99) groups. (**C** and **D**) Cumulative incidences of overall cGVHD (**C**) and moderate to severe cGVHD (**D**) in the 12 pm (n = 99) and the 6 pm (n = 99) groups. (**E**) Probabilities of GRFS in the 12 pm (n = 99) and 6 pm (n = 99) groups. Data were analyzed using Gray’s test (**A-D**) and log-rank test (**E**).

**Table 2.** Multivariate analyses for aGVHD in both the intention-to-treat and per-protocol sets.

| ITT | Multivariate analysis |  |  | PPS | Multivariate analysis |  |  |
| --- | --- | --- | --- | --- | --- | --- | --- |
|  | HR | 95%CI | P |  | HR | 95%CI | P |
| **Grade II-IV aGVHD** |  |  |  | **Grade II-IV aGVHD** |  |  |  |
| Infusion group |  |  |  | Infusion group |  |  |  |
| 12 pm | 1 |  |  | 12 pm | 1 |  |  |
| 6 pm | 2.39 | 1.13-5.04 | **0.023** | 6 pm | 3.60 | 1.42-9.17 | **0.007** |
| Disease status* | 1.67 | 1.21-2.30 | **0.002** | Disease status* | 1.81 | 1.27-2.57 | **0.001** |
| **Grade III-IV aGVHD** |  |  |  | **Grade III-IV aGVHD** |  |  |  |
| Infusion group |  |  |  | Infusion group |  |  |  |
| 12 pm | 1 |  |  | 12 pm | 1 |  |  |
| 6 pm | 6.25 | 1.39-28.15 | **0.017** | 6 pm | 10.73 | 1.30-88.37 | **0.027** |
|  |  |  |  | Disease status* | 1.87 | 1.16-3.03 | **0.011** |

Multivariate analysis of aGVHD was performed via Fine-Gray proportional hazards regression for competing events. Independent risk factors were identified via the stepwise selection method based on the BIC. All tests were two-sided, and exact *P* values are reported. \*Disease status was categorized as complete remission with negative measurable residual disease (MRD), complete remission with positive MRD, partial remission, or nonremission. MRD status was assessed within 28 days prior to transplantation using multicolor flow cytometry (MFC) (with a sensitivity of 10<sup>-4</sup> nucleated cells) and/or real-time quantitative polymerase chain reaction assays (with a sensitivity of 10<sup>-3</sup> ABL level)

### Other End Points

#### Components of the primary end point

In the intention-to-treat population, the cumulative incidence of grade III-IV aGVHD was significantly lower in the 12:00 pm group than in the 6:00 pm group (2.0% [95% CI, 0.4%-6.5%] vs 12.2% [95% CI, 6.7%-19.5%], *P* = 0.006; hazard ratio, 6.25 [95% CI, 1.39-28.15]; Fig. 2B). Similar findings were observed in the per-protocol set (12 pm vs 6 pm: 1.5% [95% CI, 0.1%-7.1%] vs 13.7% [95% CI, 7.5%-21.9%], *P* = 0.007; hazard ratio, 9.65 [95% CI, 1.24-75.09]; Fig. S2B). Multivariable analysis also identified later infusion time as an independent risk factor for grade III-IV aGVHD (hazard ratio, 6.25, [95% CI, 1.39-28.15]; *P* = 0.017) (Table 2).

#### cGVHD, engraftment and other complications

In the intention-to-treat population, the cumulative incidence of mild to severe cGVHD at 360 days tended to be lower in the 12:00 pm group than in the 6:00 pm group (27.5% [95% CI, 19.1%-36.7%] vs 38.5% [95% CI, 28.8%-48.0%], *P* = 0.107). Moreover, the cumulative incidence of moderate to severe cGVHD was significantly lower in the 12:00 pm group than in the 6:00 pm group (12.2% [95% CI, 6.6%-19.5%] vs 25.3% [95% CI, 17.2%-34.2%]; *P* = 0.018) (Figs. 2C and 2D). The trend was consistent in the per-protocol set (Figs. S2C and S2D). The distribution of cGVHD severity and organ involvement between the two groups are shown in Table S6.

Neutrophil engraftment occurred at a median of 13 days in the 12:00 pm group and 12 days in the 6:00 pm group. Platelet recovery occurred at a median of 16 days in both the 12:00 pm and 6:00 pm groups. No significant between-group differences were observed in the cumulative incidence of neutrophil engraftment or platelet recovery in either the intention-to-treat or per-protocol populations (Table 3 and Table S7). The incidence of other transplantation-related complications, including cytomegalovirus and Epstein-Barr virus reactivation, bacterial infection, engraftment syndrome, hepatic veno-occlusive disease, and hemorrhagic cystitis, did not differ significantly between groups (Table S8).

**Table 3.**
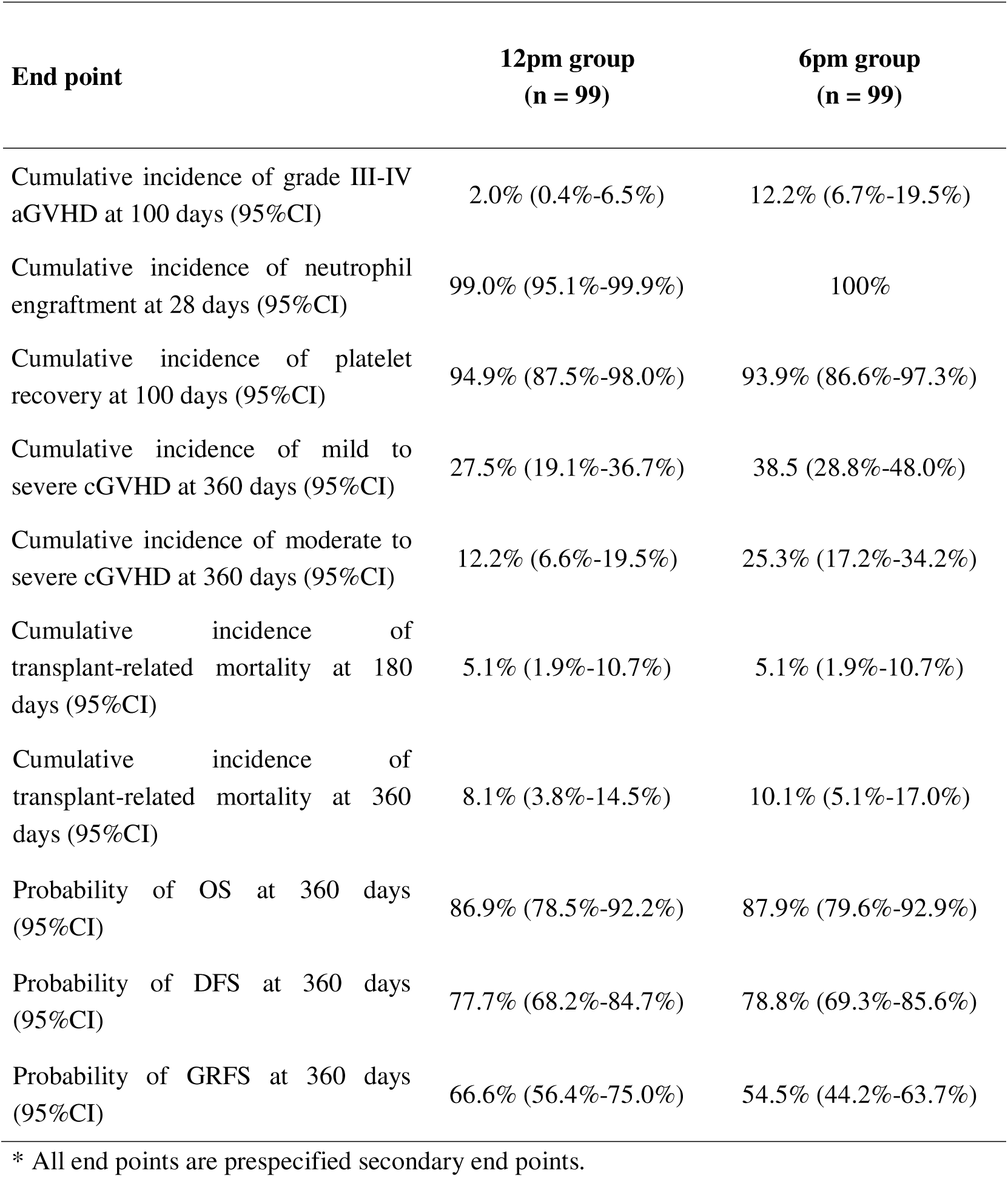
Secondary end points in the intention-to-treat population*.

#### Survival

No significant between-group differences were observed in TRM at either 180 or 360 days, relapse, OS, or DFS at 360 days (Table 3 and Table S7). A total of 14 patients in the 12:00 pm group and 15 patients in the 6:00 pm group died during follow-up; the causes of death are shown in Table S9. The median GRFS in the intention-to-treat population at 360 days was longer in the 12:00 pm group than in the 6:00 pm group (17.0 months vs. 15.1 months, hazard ratio, 1.56 [95% CI, 0.99-2.44]) (Fig. 2E). Follow-up for secondary end points is ongoing, and these findings should be interpreted with caution.

## Discussion

In this multicenter, phase 3 RCT, the primary end point was achieved, demonstrating that stem cell infusion at 12:00 pm was associated with significantly lower incidences of grade II-IV and III-IV aGVHD than infusion at 6:00 pm. Notably, despite the reduction in aGVHD risk, there were no differences between groups in relapse, hematopoietic recovery, infectious events, or other transplantation-related complications. The 12:00 pm group exhibited a lower incidence of cGVHD, whereas GRFS was numerically higher but did not reach statistical significance.

Differential adherence to the assigned infusion window should be considered when interpreting the results. More deviations occurred in the 12:00 pm group, largely because completing stem cell collection, processing, and preparation before noon was more difficult than scheduling infusion later in the day. These deviations were mainly related to the transplantation workflow rather than to patients’ clinical condition. Because most deviations reduced the difference in actual infusion time between the 2 groups, they would be more likely to attenuate than exaggerate the treatment effect. Similar results in the intention-to-treat and per-protocol analyses provide further reassurance that the primary finding was not driven by protocol deviations.

The selection of infusion time points in this trial was guided by prior clinical observations and practical clinical considerations. In our previous retrospective analyses, stem cell infusion before 2:00 pm was associated with a lower incidence and severity of aGVHD.^14^ Given that peripheral blood stem cell collection typically takes 3 to 4 hours and is initiated in the morning, 12:00 pm was chosen as an early, clinically feasible infusion time. In contrast, 6:00 pm was selected as a later time point that aligns routine clinical workflows, including cell processing, nursing coordination, and patient preparation.

Optimal infusion timing may differ across other types of allo-HSCT, such as cord blood transplantation and unrelated donor transplantation. For thawed cord blood grafts, the omission of on-site stem cell collection substantially relieves time constraints, potentially enabling earlier infusion, as supported by our retrospective findings.^15^ We have accordingly launched a dedicated RCT to validate the time-of-day effect in cord blood transplantation (NCT07047456). By comparison, stem cell products from unrelated donors often require long-distance transportation, introducing additional clinical uncertainty. In this setting, infusion scheduling needs to balance the circadian impact on aGVHD risk alongside logistical concerns related to stem cell preservation and cell viability during transit and waiting time before infusion.

This study has several limitations. First, adherence to the prespecified infusion time window differed between groups, with a higher proportion of protocol deviations in the 12:00 pm group. Although the intention-to-treat analysis preserves the benefits of randomization and the per-protocol analysis yielded consistent findings, differential adherence may have influenced the estimated treatment effect. Second, the trial was open-label, and GVHD events were not adjudicated by an independent blinded central committee. However, aGVHD was assessed according to standardized MAGIC criteria, and each diagnosis was confirmed by at least 2 transplant physicians with supporting clinical data recorded in the EDC system. Nevertheless, some risk of ascertainment or grading bias remains. Third, only a small number of patients underwent matched unrelated donor transplantation or received PT-Cy–based GVHD prophylaxis without ATG. Therefore, the generalizability of these findings to transplantation settings in which matched unrelated donors or PT-Cy–based prophylaxis are more commonly used may be limited. Finally, although follow-up was sufficient for all prespecified 360-day end points, longer-term follow-up is needed to determine whether infusion timing influences late cGVHD and long-term survival outcomes.

Despite these limitations, the present RCT provides robust prospective evidence that stem cell infusion timing significantly affects aGVHD risk. To date, evidence linking infusion timing to transplantation outcomes has been limited to retrospective analyses and preclinical studies, which are inherently subject to confounding and variability in treatment delivery.

From a clinical perspective, stem cell infusion timing represents a feasible, low-cost, and readily implementable intervention that does not require modification of existing allo-PBSCT protocols. Unlike pharmacologic strategies for GVHD prevention, which may increase the risk of relapse or infection, ^21,22^ optimization of infusion timing may reduce aGVHD risk without compromising immune reconstitution or disease control. Collectively, these findings suggest that integrating circadian biology considerations into allo-PBSCT practice may may provide a practical approach to reducing aGVHD risk.

## Supporting information

Data S1-Protocol

## Data Availability

All data produced in the present study are available upon reasonable request to the authors.

## Acknowledgments

We are deeply grateful to all the patients and their families who participated in this study. We also thank the clinical staff, including physicians, nurses, and technical personnel, for their contributions to stem cell collection, processing, and infusion. Xiaoyu Zhu is supported by grants from the National Key Research and Development Program (2025YFA1804700), the National Natural Science Foundation of China (U23A20453, 82270223), the USTC Research Funds of the Double First-Class Initiative (YD9110002047), the Anhui Provincial Department of Education Scientific Research Project (2023AH010079) and the Anhui Provincial Natural Science Foundation (2308085J09). Cheng Zhan is supported by grants from the National Natural Science Foundation of China (32525034, 82495183, 32271063) and the Strategic Priority Research Program of the Chinese Academy of Sciences (grant no. XDB0940000). Yue Wu is supported by grants from the National Natural Science Foundation of China (82600471) and the USTC Research Funds of the Double First-Class Initiative (YD9115202642).

## Author Contributions

Xiaoyu Zhu and Cheng Zhan had full access to all the data in the study and take responsibility for the integrity of the data and the accuracy of the data analysis. Xiaoyu Zhu and Cheng Zhan conceived and designed the study. All authors contributed to the acquisition, analysis, or interpretation of the data. Yue Wu performed the statistical analyses under the statistical supervision of Weiwei Wu. Yue Wu, Xiaoyu Zhu, and Cheng Zhan drafted the manuscript, and all authors critically revised it for important intellectual content. Yang Yang, Xiaoxia Hu, Weijie Cao, Yanmin Zhao, Yujun Dong, Yang Cao, Jiayu Huang, Ran Zhang, Kaidi Song, Guangyu Sun, and Wen Yao were responsible for patient management and clinical outcome assessment. Qing Cheng and Jian Wang were responsible for stem-cell collection and infusion procedures. All authors contributed to data curation and oversight and provided administrative, technical, or material support. Xiaoyu Zhu supervised the study. All authors reviewed and approved the final manuscript.

## Data sharing statement

Deidentified individual participant data underlying the results reported in this article will be available to qualified researchers through the study EDC system upon reasonable request to the corresponding author and approval of the proposed use.

