## Supplementary Appendix for "Effect of stem cell infusion timing on acute graft-versus-host disease: a randomized clinical trial"

This appendix has been provided by the authors to give readers additional information about their work.

Updated Sep 1, 2026

### Section A. Infusion time & aGVHD-001 Investigational Team

**Primary Investigator***

| **Primary Investigator** | **Site** | **Location** | **Patients enrolled** |
| --- | --- | --- | --- |
| **Xiaoyu Zhu** | **The First Affiliated Hospital of University of Science and Technology of China** | **Hefei, Anhui, China** | **33** |
| **Xiaoxia Hu** | **Rui Jin Hospital Affiliated to Shanghai Jiao Tong University** | **Shanghai, Shanghai, China** | **79** |
| **Yang Cao** | **Tongji Hospital, Tongji Medical College, Huazhong University of Science and**  **Technology** | **Wuhan, Hubei, China** | **49** |
| **Yanmin Zhao** | **The First Affiliated Hospital, Zhejiang University School of Medicine** | **Hangzhou, Zhejiang, China** | **8** |
| **Yujun Dong** | **Peking University First Hospital** | **Beijing, Beijing, China** | **8** |
| **Weijie Cao** | **The First Affiliated Hospital of Zhengzhou University** | **Zhengzhou, Henan, China** | **21** |

***Primary investigators who screened and/or randomized participants.**

### Section B. GVHD Prophylaxis Regimens

Calcineurin inhibitors were initiated before transplantation (day -1) and adjusted to target therapeutic levels, followed by tapering beginning at approximately day +60 and discontinuation by day +100 in the absence of GVHD. MMF was administered from day +1 to approximately day +40. The administration of rATG, ATLG, PT-Cy and MTX complied with institutional protocols. rATG was administered at a total dose of 4.5-7.5 mg/kg from day -4 to -2 or 2.5 mg/kg on day +15 or +16 post transplantation; ATLG was administered at a total dose of 10-25 mg/kg from day -4 to -2. PT-Cy was administered on days +3 and +4 at a total dose of 60-100 mg/kg. MTX was administered intravenously on day +1 at 15 mg/m2 and 10 mg/m2 on days +3 and +6 post transplantation.

### Section C. Statistical Methods

Time-to-event end points involving competing risks were analyzed using cumulative incidence functions and compared between groups using Gray’s test.^1^ For aGVHD and cGVHD, death and relapse without prior GVHD were treated as competing events. For TRM and relapse, each was considered a competing risk for the other. For neutrophil engraftment and platelet recovery, death before engraftment was treated as a competing event. OS, DFS, and GRFS were estimated using the Kaplan-Meier method and compared using the log-rank test. All authors vouch for the completeness and accuracy of the data and for the fidelity of the trial to the protocol.

Prespecified multivariable analyses were performed with adjustment for clinically relevant baseline and transplantation-related characteristics. Prespecified covariates included age, sex, disease diagnosis, disease status at transplantation, conditioning intensity (MAC vs. RIC), donor types, infused CD34^+^ cell dose, donor-recipient HLA compatibility, donor-recipient sex compatibility (female donor to male recipient vs. others), GVHD prophylaxis regimen, and methotrexate use. These analyses were performed using Fine-Gray sub-distribution hazard models.^2,3^ The proportional subdistribution hazards assumption for the Fine-Gray models was assessed by testing time-dependent interactions between covariates and follow-up time. There were no missing data for the variables included in the primary and secondary outcome analyses. The time-to-event analyses were conducted using censoring at the last available follow-up. All statistical tests were 2-sided, with a significance level of 0.05. Statistical analyses were performed using R software (version 4.2.2)^4^.

### Figure S1. Distribution of stem cell infusion start times. Shaded areas represent the per-protocol population in the 12 pm and 6 pm groups.

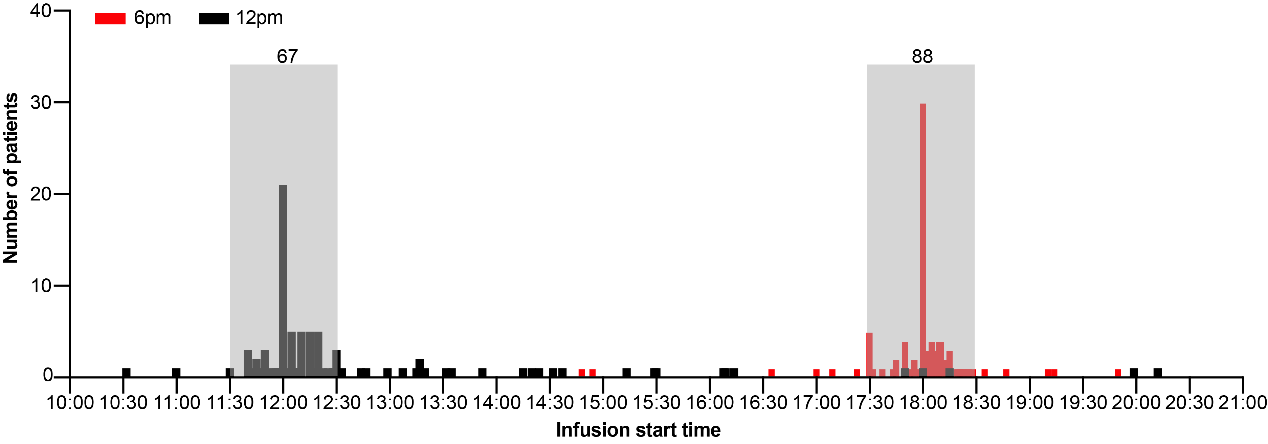

### Figure S2. Effects of stem cell infusion timing on GVHD and survival in the per-protocol set.

(**A** and **B**) Cumulative incidences (*CI*) of grade II-IV aGVHD (**A**) and grade III-IV aGVHD (**B**) in the 12 pm (n = 67) and 6 pm (n = 88) groups.

(**C** and **D**) Cumulative incidences of overall cGVHD (**C**) and moderate to severe cGVHD (**D**) in the 12 pm (n = 67) and 6 pm (n = 88) groups.

(**E**) Probabilities of GRFS in the 12 pm (n = 67) and 6 pm (n = 88) groups.

Data were analyzed using Gray's test (**A-D**) and log-rank test (**E**).

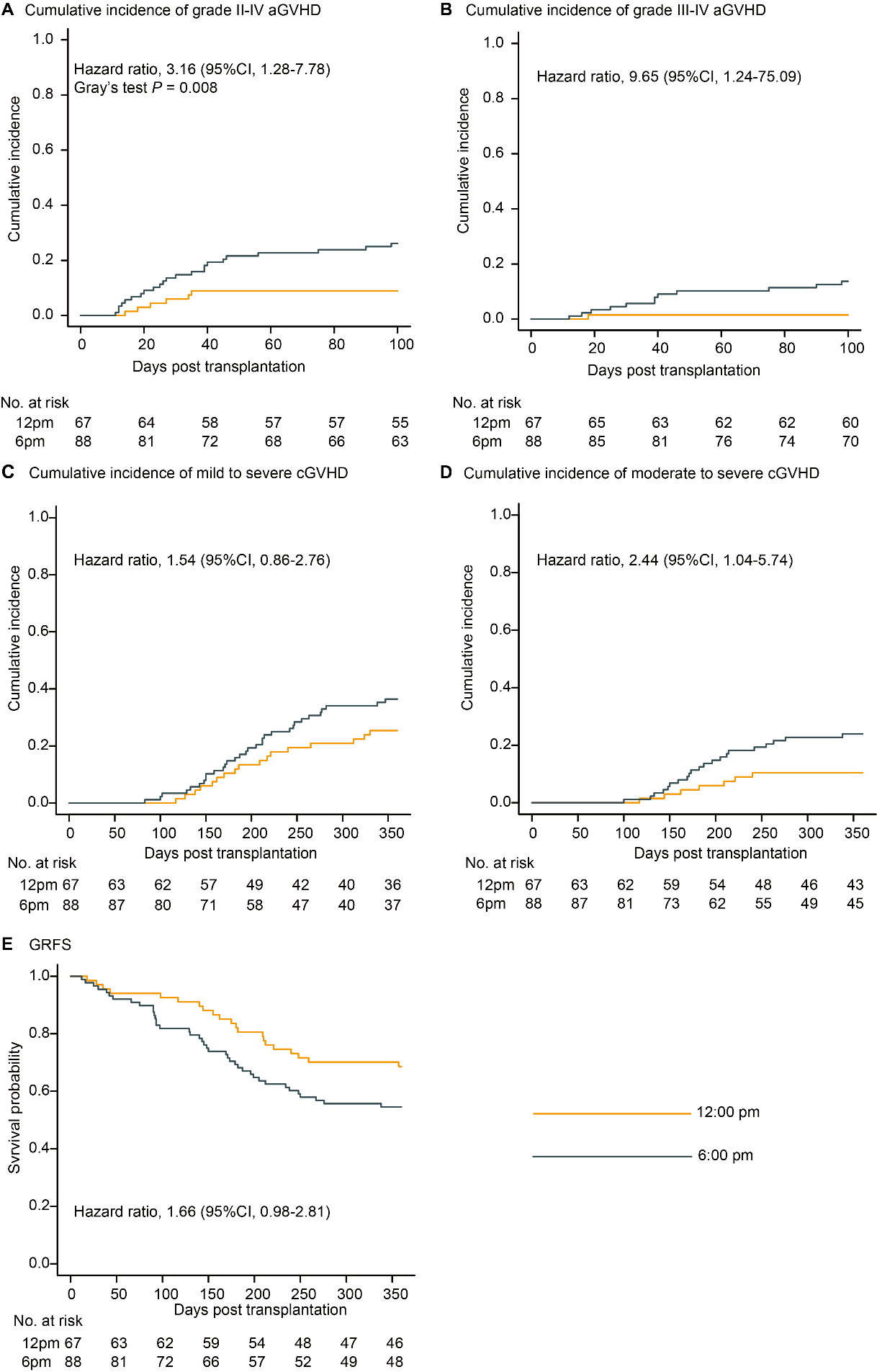

### Table S1. Details of conditioning regimen in the intention-to-treat population.

| Characteristics | 12pm group  (n = 99) | 6pm group  (n = 99) |
| --- | --- | --- |
| Conditioning regimen (n) |  |  |
| Bu-based |  |  |
| Flu (150 mg/m^2^) + Bu (6.4 mg/kg) + Mel (100 mg/m^2^) ± VP16 (400 mg/m^2^) | 31 | 40 |
| Bu (9.6-12.8 mg/kg) +Flu (120-150 mg/m^2^) + Cy (14.5 mg/kg or 1.8 g/m^2^ ×2d) / Mel (100-120 mg/m^2^) ± Ara-C (2 g/m^2^ ×4d) / Dec (20 mg/m^2^ ×3d)/CCNU (250 mg/m^2^) | 60 | 53 |
| Irradiation-based |  |  |
| TBI (12 Gy) + Cy (120 mg/kg) + CCNU (250 mg/m^2^) | 6 | 3 |
| TMI (15 Gy) + Cy (80-120 mg/kg) | 2 | 3 |

Flu fludarabine, Bu busulfan, Cy cyclophosphamide, Mel melphalan, VP16 Etoposide, Ara-C cytarabine, Dec decitabine, CCNU simustine, TBI total body irradiation, TMI total marrow irradiation

### Table S2. Baseline Characteristics of Patients in the Per-Protocol Population.

| Characteristics | 12pm group  (n = 67) | 6pm group  (n = 88) | Total  (n = 155) |
| --- | --- | --- | --- |
| Sex, n (%) |  |  |  |
| Male | 39 (58.2) | 58 (65.9) | 97 (62.6) |
| Female | 28 (41.8) | 30 (34.1) | 58 (37.4) |
| Age (years), Median (Range) | 38 (14-60) | 39 (13-60) | 38 (13-60) |
| Weight(kg), Median (Range) | 63.3 (40.3-104.1) | 63.5 (43.6-107.9) | 63.3 (40.3-107.9) |
| Diagnostics, n (%) |  |  |  |
| AML | 28 (41.8) | 47 (53.4) | 75 (48.4) |
| ALL | 20 (29.9) | 22 (25.0) | 42 (27.1) |
| MDS/MPN | 15 (22.4) | 12 (13.6) | 27 (17.4) |
| Others^#^ | 4 (6.0) | 7 (8.0) | 11 (7.1) |
| Disease status*, n (%) |  |  |  |
| CR/CRi^MRD-^ | 44 (65.7) | 59 (67.0) | 103 (66.5) |
| CR^MRD+^ | 15 (22.4) | 21 (23.9) | 36 (23.2) |
| PR | 4 (6.0) | 3 (3.4) | 7 (4.5) |
| NR | 4 (6.0) | 5 (5.7) | 9 (5.8) |
| CD34^+^cell infused（10^6^/kg） | 6.73 (2.19-43.77) | 6.40 (3.16-10.59) | 6.57 (2.19-43.77) |
| TNCs infused（10^8^/kg） | 10.08 (3.00-33.39) | 9.03 (1.32-63.98) | 9.66 (1.32-63.98) |
| Female to male, n (%) |  |  |  |
| Yes | 8 (11.9) | 17 (19.3) | 25 (16.1) |
| No | 59 (88.1) | 71 (80.7) | 130 (83.9) |
| HLA compatibility (/10), n (%) |  |  |  |
| 10 | 10 (14.9) | 14 (15.9) | 24 (15.5) |
| 8-9 | 5 (7.5) | 17 (19.3) | 22 (14.2) |
| 6-7 | 17 (25.4) | 27 (30.7) | 44 (28.4) |
| 5 | 35 (52.2) | 30 (34.1) | 65 (41.9) |
| aGVHD prophylaxis regimen, n (%) |  |  |  |
| CNIs+MMF without ATG or PT-Cy | 3 (4.5) | 5 (5.7) | 8 (5.2) |
| CNIs+MMF with ATG | 58 (86.6) | 64 (72.7) | 122 (78.7) |
| CNIs+MMF with PT-cy | 1 (1.5) | 2 (2.3) | 3 (1.9) |
| CNIs+MMF with ATG and PT-Cy | 5 (7.5) | 17 (19.3) | 22 (14.2) |
| MTX prophylaxis, n (%) | 53 (79.1) | 64 (72.7) | 117 (75.5) |
| ABO incompatibility, n (%) |  |  |  |
| Identical | 39 (58.2) | 42 (47.7) | 81 (52.3) |
| Minor incompatibility | 18 (26.9) | 20 (22.7) | 38 (24.5) |
| Major incompatibility | 8 (11.9) | 18 (20.5) | 26 (16.8) |
| Bidirectional incompatibility | 2 (3.0) | 8 (9.1) | 10 (6.5) |
| Conditioning regimen, n (%) |  |  | 0 |
| MAC | 45 (67.2) | 54 (61.4) | 99 (63.9) |
| RIC | 22 (32.8) | 34 (38.6) | 56 (36.1) |
| Donor type, n (%) |  |  |  |
| Haplo-identical | 57 (85.1) | 73 (83.0) | 130 (83.9) |
| MSD | 10 (14.9) | 14 (15.9) | 24 (15.5) |
| URD | 0 (0.0) | 1 (1.1) | 1 (0.6) |
| Follow-up time among survivors (d) | 685(404-834) | 650 (403-834) | 655 (403-834) |

CNIs calcineurin inhibitors, MMF mycophenolate mofetil, ATG antithymocyte globulin, PT-Cy post-transplant cyclophosphamide, MTX methotrexate, MAC myeloablative conditioning regimen, RIC reduced intensity conditioning, TNCs total nucleated cells, MSD matched sibling donor, URD unrelated donor. *MRD status was assessed separately for AML and ALL patients within 28 days prior to transplantation using real-time quantitative polymerase chain reaction assays (with a sensitivity of 10⁻⁴ ABL level) and/or multicolor flow cytometry (MFC) (with a sensitivity of 10⁻⁴ nucleated cells); ^#^MPAL, mixed phenotype acute leukemia; NHL, non-Hodgkin's lymphoma; BPDCN, blastic plasmacytoid dendritic cell neoplasm.

### Table S3. Baseline characteristics according to adherence to the assigned infusion time window

| Characteristics | 12pm group | | | 6pm group | | | | |
| --- | --- | --- | --- | --- | --- | --- | --- | --- |
|  | Compliant (n = 67) | Noncompliant (n = 32) | *P*  value | Compliant (n = 88) | Noncompliant (n = 11) | | | *P*  value |
| Sex, n (%) |  |  | 0.669 |  | |  | 0.201 | |
| Male | 39 (58.2) | 17 (53.1) |  | 58 (65.9) | | 5 (45.5) |  | |
| Female | 28 (41.8) | 15 (46.9) |  | 30 (34.1) | | 6 (54.5) |  | |
| Age (years), Median (Range) | 38 (14-60) | 37 (14-59) | 0.122 | 39 (13-60) | | 36 (17-59) | 0.518 | |
| Weight(kg), Median (Range) | 63.3 (40.3- 104.1) | 62.0 (39.4-88.9) | 0.503 | 63.5 (43.6- 107.9) | | 59.5 (50.1-70.0) | 0.130 | |
| Diagnostics, n (%) |  |  | 0.184 |  | |  | 0.842 | |
| AML | 28 (41.8) | 18 (56.2) |  | 47 (53.4) | | 6 (54.5) |  | |
| ALL | 20 (29.9) | 11 (34.4) |  | 22 (25.0) | | 4 (36.4) |  | |
| MDS/MPN | 15 (22.4) | 2 (6.2) |  | 12 (13.6) | | 1 (9.1) |  | |
| Others^#^ | 4 (6.0) | 1 (3.1) |  | 7 (8.0) | | 0 (0.0) |  | |
| Disease status*, n (%) |  |  | 0.569 |  | |  | 0.260 | |
| CR/CRi^MRD-^ | 44 (65.7) | 24 (75.0) |  | 59 (67.0) | | 9 (81.8) |  | |
| CR^MRD+^ | 15 (22.4) | 6 (18.8) |  | 21 (23.9) | | 1 (9.1) |  | |
| PR | 4 (6.0) | 0 (0.0) |  | 3 (3.4) | | 1 (9.1) |  | |
| NR | 4 (6.0) | 2 (6.2) |  | 5 (5.7) | | 0 (0.0) |  | |
| CD34^+^cell infused（10^6^/kg） | 6.73 (2.19-43.77) | 6.25 (3.40-9.99) | 0.115 | 6.70 (3.16-10.59) | | 6.36 (4.80-9.70) | 0.570 | |
| TNCs infused（10^8^/kg） | 10.08 (3.00- 33.39) | 8.29 (3.20-31.40) | 0.206 | 9.03 (1.32- 63.98) | | 9.39 (3.80-17.57) | 0.867 | |
| Female to male, n (%) |  |  | 0.751 |  | |  | 0.683 | |
| Yes | 8 (11.9) | 5 (15.6) |  | 17 (19.3) | | 1 (9.1) |  | |
| No | 59 (88.1) | 27 (84.4) |  | 71 (80.7) | | 10 (90.9) |  | |
| HLA compatibility (/10), n (%) |  |  | 0.486 |  | |  | 0.335 | |
| 10 | 10 (14.9) | 5 (15.6) |  | 14 (15.9) | | 4 (36.4) |  | |
| 8-9 | 11 (16.4) | 9 (28.1) |  | 17 (19.3) | | 2 (18.2) |  | |
| 6-7 | 17 (25.4) | 10 (31.2) |  | 27 (30.7) | | 4 (36.4) |  | |
| 5 | 29 (43.3) | 8 (25.0) |  | 30 (34.1) | | 1 (9.1) |  | |
| aGVHD prophylaxis regimen, n (%) |  |  | 0.582 |  | |  | 0.301 | |
| CNIs+MMF without ATG or PT-Cy | 3 (4.5) | 2 (6.2) |  | 5 (5.7) | | 2 (18.2) |  | |
| CNIs+MMF with ATG | 58 (86.6) | 25 (78.1) |  | 64 (72.7) | | 6 (54.5) |  | |
| CNIs+MMF with PT-cy | 1 (1.5) | 1 (3.1) |  | 2 (2.3) | | 0 (0.0) |  | |
| CNIs+MMF with ATG and PT-Cy | 5 (7.5) | 4 (12.5) |  | 17 (19.3) | | 3 (27.3) |  | |
| MTX prophylaxis, n (%) | 53 (79.1) | 28 (87.5) | 0.409 | 64 (72.7) | | 8 (72.7) | 1.000 | |
| ABO incompatibility, n (%) |  |  | 0.695 |  | |  | 0.251 | |
| Identical | 39 (58.2) | 16 (50.0) |  | 42 (47.7) | | 7 (63.6) |  | |
| Minor incompatibility | 18 (26.9) | 9 (28.1) |  | 20 (22.7) | | 1 (9.1) |  | |
| Major incompatibility | 8 (11.9) | 3 (9.4) |  | 18 (20.5) | | 2 (18.2) |  | |
| Bidirectional incompatibility | 2 (3.0) | 4 (12.5) |  | 8 (9.1) | | 1 (9.1) |  | |
| Conditioning regimen, n (%) |  |  | 0.817 |  | |  | 0.344 | |
| MAC | 45 (67.2) | 23 (71.9) |  | 54 (61.4) | | 5 (45.5) |  | |
| RIC | 22 (32.8) | 9 (28.1) |  | 34 (38.6) | | 6 (54.5) |  | |
| Donor type, n (%) |  |  | 0.485 |  | |  | 0.103 | |
| Haplo-identical | 57 (85.1) | 26 (81.2) |  | 73 (83.0) | | 7 (63.6) |  | |
| MSD | 10 (14.9) | 5 (15.6) |  | 14 (15.9) | | 3 (27.3) |  | |
| URD | 0 (0.0) | 1 (3.1) |  | 1 (1.1) | | 1 (9.1) |  | |
| Follow-up time among survivors (d) | 644 (461-833) | 685 (404-834) | 0.324 | 697 (536-812) | | 650 (403-834) | 0.168 | |

CNIs calcineurin inhibitors, MMF mycophenolate mofetil, ATG antithymocyte globulin, PT-Cy post-transplant cyclophosphamide, MTX methotrexate, MAC myeloablative conditioning regimen, RIC reduced intensity conditioning, TNCs total nucleated cells, MSD matched sibling donor, URD unrelated donor. *MRD status was assessed separately for AML and ALL patients within 28 days prior to transplantation using real-time quantitative polymerase chain reaction assays (with a sensitivity of 10⁻⁴ ABL level) and/or multicolor flow cytometry (MFC) (with a sensitivity of 10⁻⁴ nucleated cells); ^#^MPAL, mixed phenotype acute leukemia; NHL, non-Hodgkin's lymphoma; BPDCN, blastic plasmacytoid dendritic cell neoplasm.

### Table S4. Sensitivity analysis for aGVHD with Fine-Gray proportional hazards regression, applied to varying infusion time cutoffs.

|  | Grade II-IV aGVHD HR_adjusted_ | *P*  value | Grade III-IV aGVHD HR_adjusted_ | *P* value |
| --- | --- | --- | --- | --- |
| Infusion before 1:00 pm versus after 1:00 pm | 2.77 (95% *CI*, 1.17-6.57) | 0.021 | 10.22 (95% *CI*, 1.06-98.25) | 0.044 |
| Infusion before 2:00 pm versus after 2:00 pm | 2.02 (95% *CI*, 0.96-4.25) | 0.065 | 5.05 (95% *CI*, 0.94-27.13) | 0.059 |
| Infusion before 3:00 pm versus after 3:00 pm | 2.32 (95% *CI*, 1.07-5.03) | 0.032 | 6.26 (95% *CI*, 0.96 – 40.88) | 0.055 |
| Infusion before 4:00 pm versus after 4:00 pm | 2.10 (95% *CI*, 1.00-4.43) | 0.051 | 6.40 (95% *CI*, 1.04-39.55) | 0.046 |
| Infusion before 5:00 pm versus after 5:00 pm | 2.31 (95% *CI*, 1.10-4.83) | 0.026 | 7.82 (95% *CI*, 1.18-51.78) | 0.033 |

### Table S5. Maximum aGVHD by day 100 in the intention-to-treat population.

| Characteristics | 12pm group (n = 99) | 6pm group  (n = 99) | Total  (n = 198) |
| --- | --- | --- | --- |
| Maximum severity of aGVHD, n (%) |  |  |  |
| Grade I | 5 (5.1) | 2 (2.0) | 7 (3.5) |
| Grade II | 9 (9.1) | 10 (10.1) | 19 (9.6) |
| Grade III | 2 (2.0) | 2 (2.0) | 4 (2.0) |
| Grade IV | 0 | 10 (10.1) | 10 (5.1) |
| Organ involvement in grade II-IV aGVHD patient |  |  |  |
| Skin | 9 (9.1) | 8 (8.1) | 17 (8.6) |
| Upper GI | 3 (3.0) | 12 (12.1) | 15 (7.6) |
| Lower GI | 4 (4.0) | 11 (11.1) | 15 (7.6) |
| Liver | 1 (1.0) | 3 (3.0) | 4 (2.0) |
| Maximum severity of skin in grade II-IV aGVHD patient, n (%) |  |  |  |
| Stage 0 | 2 (2.0) | 14 (14.1) | 16 (8.1) |
| Stage 1 | 1 (1.0) | 2 (2.0) | 3 (1.5) |
| Stage 2 | 3 (3.0) | 2 (2.0) | 5 (2.5) |
| Stage 3 | 5 (5.1) | 4 (4.0) | 9 (4.5) |
| Maximum severity of upper GI in grade II-IV aGVHD patient, n (%) |  |  |  |
| Stage 0 | 8 (8.1) | 10 (10.1) | 18 (9.1) |
| Stage 1 | 3 (3.0) | 12 (12.1) | 15 (7.6) |
| Maximum severity of lower GI in grade II-IV aGVHD patient, n (%) |  |  |  |
| Stage 0 | 7 (7.1) | 11 (11.1) | 18 (9.1) |
| Stage 1 | 3 (3.0) | 2 (2.0) | 5 (2.5) |
| Stage 2 | 0 | 1 (1.0) | 1 (0.5) |
| Stage 3 | 1 (1.0) | 0 | 1 (0.5) |
| Stage 4 | 0 | 8 (8.1) | 8 (4.0) |
| Maximum severity of liver in grade II-IV aGVHD patient, n (%) |  |  |  |
| Stage 0 | 10 (10.1) | 19 (19.2) | 29 (14.6) |
| Stage 1 | 1 (1.0) | 2 (2.0) | 3 (1.5) |
| Stage 2 | 0 | 1 (1.0) | 1 (0.5) |

GI gastrointestinal tract

### Table S6. Maximum cGVHD by day 360 in the intention-to-treat population.

| Characteristics | 12pm group (n = 99) | 6pm group (n = 99) | Total  (n = 198) |
| --- | --- | --- | --- |
| Maximum severity of cGVHD, n (%) |  |  |  |
| None | 69 (69.7) | 59 (59.6) | 128 (64.6) |
| Mild | 17 (17.2) | 15 (15.2) | 32 (16.2) |
| Moderate | 8 (8.1) | 19 (19.2) | 27 (13.6) |
| Severe | 5 (5.1) | 6 (6.1) | 11 (5.6) |
| Organ involvement in cGVHD patient |  |  |  |
| Skin | 19 (19.2) | 21 (21.2) | 40 (20.2) |
| GI | 2 (2.0) | 3 (3.0) | 5 (2.5) |
| Mouth | 8 (8.1) | 12 (12.1) | 20 (10.1) |
| Liver | 12 (12.1) | 9 (9.1) | 21 (10.6) |
| Eye | 8 (8.1) | 4 (4.0) | 12 (6.1) |
| Lung | 4 (4.0) | 6 (6.1) | 10 (5.1) |
| Joints and fascia | 1 (1.0) | 5 (5.1) | 6 (3.0) |
| Genital tract | 1 (1.0) | 1 (1.0) | 2 (1.0) |

GI gastrointestinal tract

### Table S7. Second end points in the per-protocol set*.

| **End point** | **12pm group**  **(n = 67)** | **6pm group**  **(n = 88)** |
| --- | --- | --- |
| Cumulative incidence of grade III-IV aGVHD at 100 days (95%CI) | 1.5% (0.1%-7.1%) | 13.7% (7.5%-21.9%) |
| Cumulative incidence of neutrophil engraftment at 28 days (95%CI) | 98.5% (92.9%-99.9%) | 100% |
| Cumulative incidence of platelet recovery at 100 days (95%CI) | 94.0% (83.2%-98.0%) | 93.2% (885.0%-97.0%) |
| Cumulative incidence of mild to severe cGVHD at 360 days (95%CI) | 25.4% (15.6%-36.3%) | 36.4% (26.4%-46.5%) |
| Cumulative incidence of moderate to severe cGVHD at 360 days (95%CI) | 10.4% (4.6%-19.2%) | 23.9% (15.5%-33.3%) |
| Cumulative incidence of transplant-related mortality at 180 days (95%CI) | 6.0% (1.9%-13.4%) | 5.7% (2.1%-11.9%) |
| Cumulative incidence of transplant-related mortality at 360 days (95%CI) | 10.4% (4.6%-19.2%) | 11.4% (5.8%-19.0%) |
| Probability of OS at 360 days (95%CI) | 85.1% (74.0%-91.7%) | 86.4% (77.2%-92.0%) |
| Probability of DFS at 360 days (95%CI) | 79.1% (67.2%-87.0%) | 77.3% (67.0%-84.7%) |
| Probability of GRFS at 360 days (95%CI) | 68.6% (56.0%-78.3%) | 54.5% (43.6%-64.2%) |

* All end points are prespecified secondary end points.

### Table S8. Complications post transplantation in the intention-to-treat population.

| Characteristics | 12pm group  (n=99) | 6pm group  (n=99) | *P* |
| --- | --- | --- | --- |
| CMV antigenemia | 28 (28.3) | 25 (25.3) | 0.748 |
| EBV antigenemia | 32 (32.3) | 28 (28.3) | 0.643 |
| Bacterial infection | 16 (16.2) | 25 (25.3) | 0.160 |
| ES | 10 (10.1) | 10 (10.1) | 1.000 |
| HVOD | 2 (2.0) | 0 | 0.497 |
| HC | 21 (21.2) | 27 (27.3) | 0.407 |

CMV cytomegalovirus, EBV Epstein-Barr Virus, ES engraftment syndrome, HVOD, hepatic veno-occlusive disease, HC hemorrhagic cystitis

### Table S9. Details on the cause of death in the intention-to-treat population.

| Cause | 12pm group  (n = 99) | 6pm group  (n = 99) |
| --- | --- | --- |
|  | number/total number (percent) | |
| Relapse | 5/14 (35.7) | 3/15 (20.0) |
| Uncontrolled GVHD | 0 | 4/15 (26.7) |
| Infection | 5/14 (35.7) | 6/15 (40.0) |
| MODS | 3/14 (21.4) | 2/15 (13.3) |
| Engraftment failure | 1/14 (7.1) | 0 |

MODS, multi-organ dysfunction syndrome

### References.

1.	Klein JP, Rizzo JD, Zhang MJ, Keiding N. Statistical methods for the analysis and presentation of the results of bone marrow transplants. Part I: unadjusted analysis. Bone Marrow Transplant 2001;28(10):909-15. DOI: 10.1038/sj.bmt.1703260.

2.	Fine JP, Gray RJ. A proportional hazards model for the subdistribution of a competing risk. J Am Stat Assoc 1999;94:496–509.

3.	Klein JP, Rizzo JD, Zhang MJ, Keiding N. Statistical methods for the analysis and presentation of the results of bone marrow transplants. Part 2: Regression modeling. Bone Marrow Transplant 2001;28(11):1001-11. DOI: 10.1038/sj.bmt.1703271.

4.	Scrucca L, Santucci A, Aversa F. Competing risk analysis using R: an easy guide for clinicians. Bone Marrow Transplant 2007;40(4):381-7. DOI: 10.1038/sj.bmt.1705727.
