## Supplementary material for "Effect of stem cell infusion timing on acute graft-versus-host disease: a randomized clinical trial": Data S1-Protocol

**Clinical trial protocol**

Multicenter, randomized, controlled, open-label clinical study

Title A Multicenter Randomized Controlled Study of the Effect of Stem Cell Infusion Time on the Development of aGVHD in Patients With Hematological Malignancies After Allogeneic Peripheral Blood Hematopoietic Stem Cell Transplantation

Institution The First Affiliated Hospital of USTC (Anhui Provincial Hospital)

Principal investigator Xiaoyu Zhu, MD

Version and date: Version 1.0, Feb 8, 2024

1. **Study background**

Allogeneic hematopoietic stem cell transplantation (allo-HSCT) represents a pivotal therapeutic modality with the potential to cure a broad spectrum of malignant and nonmalignant hematological disorders. Allo-HSCT is widely utilized in diseases such as acute leukemia, severe aplastic anemia, congenital immunodeficiency diseases, and inherited metabolic disorders, allowing for the reconstitution of normal hematopoietic and immune function.

The world’s first hematopoietic stem cell transplantation was performed in 1957, followed by the successful first unrelated donor transplantation between two nonrelatives in 1973 and the landmark first unrelated cord blood transplantation in 1988. Since then, the number of all types of hematopoietic stem cell transplantations has witnessed exponential growth, with allo-HSCT emerging as the most prominent category. It is estimated that by 2021, a cumulative total of 20,000 hematopoietic stem cell transplantations had been completed in the United States, among which over 8,000 cases were allo-HSCT procedures. In China, starting in 2020, the annual number of allo-HSCTs has exceeded 10,000, reaching 12,632 cases in 2022 (data from the Hematopoietic Stem Cell Transplantation Group of the Hematology Branch of the Chinese Medical Association).

Despite remarkable advancements in donor selection, pretransplant conditioning regimens, and graft sources achieved in recent research, graft-versus-host disease (GVHD), particularly acute graft-versus-host disease (aGVHD), remains one of the primary factors influencing transplant-related mortality and patient quality of life. Even with effective pharmacological prophylaxis, the incidence of posttransplant aGVHD remains as high as 40-60% [1], and the mortality rate for those who develop aGVHD may reach 55%, which significantly impairs patients’ overall survival rate and quality of life. According to the 2020 statistical data from the Center for International Blood and Marrow Transplant Research (CIBMTR), in the United States, GVHD constitutes 13% and 14% of transplantation-related mortality (TRM) in HLA-identical sibling and unrelated donor transplantations, respectively. Among them, grade III-IV aGVHD is the major contributor to posttransplant mortality in patients. In 2022, the European Group for Blood and Marrow Transplantation (EBMT) conducted a retrospective analysis of 102,557 patients with hematological malignancies who underwent sibling or unrelated stem cell transplantation over different periods to investigate the changes in the incidence of aGVHD and its impact on survival outcomes. The results demonstrated that among patients who developed grade III-IV aGVHD between 2011 and 2015, the 3-year overall survival rate was merely 29% [2].

Current strategies for aGVHD prophylaxis predominantly rely on multiple immunosuppressive agents, such as calcineurin inhibitors and antimetabolites, which suppress T-cell activation in the graft. However, these pharmacological agents exert nonspecific effects, while preventing and treating aGVHD, they may increase the risk of infection and compromise the graft-versus-leukemia (GVL) effect. Moreover, these approaches overlook the impact of recipient-specific functional alterations on the graft.

Circadian rhythms, governed by the central clock, regulate immune and metabolic processes, encompassing behavioral activity, sleep-wake cycles, body temperature, metabolism, and immune responses [3]. Human physiological functions such as sleep patterns, hormone secretion, metabolism, body temperature regulation, and immune competence are all governed by the circadian clock, exhibiting rhythmic variations under both homeostatic and stress conditions. Previous studies have documented that immune functions, including peripheral blood leukocyte counts and cytokine secretion, display distinct circadian oscillations in humans. The number of mature leukocytes (excluding effector CD8^+^ T cells) in the peripheral blood peaks during the inactive phase (corresponding to nighttime in humans) and declines during the active phase (daytime in humans). Additionally, the levels of various proinflammatory cytokines in the peripheral blood, such as interferon γ (IFNγ), tumor necrosis factor-α (TNF-α), interleukin (IL)-1, IL-2, IL-6, and IL-12, also reach their zenith during the inactive phase [4].

Circadian rhythm disruption leading to immune dysregulation constitutes the pathogenic basis of numerous diseases. Meanwhile, the therapeutic efficacy of various autoimmune diseases and malignant tumors exhibits differential responses influenced by circadian rhythms. A recent clinical study investigating immune checkpoint inhibitor therapy in melanoma patients revealed that patients who received treatment before 4:30 pm demonstrated significantly prolonged long-term survival rates and superior treatment outcomes compared with those who underwent infusion after 4:30 pm [5]. This finding suggests that administering the right treatment at the optimal time window may improve patient prognosis.

Allo-HSCT is essentially a cellular therapy that reconstitutes patients’ normal hematopoietic and immune functions via the infusion of donor-derived stem cells. Whether modulating the timing of stem cell infusion can influence the incidence of aGVHD post allo-HSCT and enhance patient prognosis remains to be elucidated.

In a preliminary retrospective analysis, we evaluated clinical data from 204 patients who underwent allogeneic peripheral blood stem cell transplantation (allo-PBSCT) at the Department of Hematology, The First Affiliated Hospital of the University of Science and Technology of China (unpublished data). The cohort included 137 patients who received matched sibling donor transplantation (MSDT) and 67 patients who underwent haploidentical donor transplantation (HIDT). We investigated the association between the timing of stem cell infusion and the incidence of aGVHD. The analysis demonstrated that (1) patients who received stem cell infusion before 2:00 pm had significantly lower incidences of grade II-IV and grade III-IV aGVHD compared with those infused after 2:00 pm; (2) earlier infusion (before 2:00 pm) was associated with a significantly higher 3-year GVHD-free, relapse-free survival (GRFS); and (3) in multivariable regression analyses adjusting for potential confounders—including patient age, sex, disease status before transplantation, donor type, infused cell dose, conditioning regimen, and GVHD prophylaxis regimen—the timing of stem cell infusion remained an independent risk factor for both grade II-IV and grade III-IV aGVHD.

These findings from our single-center retrospective study suggest that the timing of stem cell infusion may significantly influence the incidence and severity of aGVHD, as well as long-term GRFS following transplantation. To date, no prospective randomized clinical trials have evaluated the effect of stem cell infusion timing on aGVHD after allo-HSCT. Given the potential for residual confounding inherent in retrospective analyses, a well-designed prospective study is warranted. Therefore, we propose a multicenter, prospective, randomized, phase 3 clinical trial to evaluate the effect of stem cell infusion timing on transplantation outcomes in patients aged 12 to 60 years with malignant hematologic diseases undergoing allo-PBSCT. Patients will be randomly assigned according to predefined infusion time points to assess whether infusion timing influences the incidence and severity of aGVHD, as well as long-term GRFS.

1) To determine whether stem cell infusion timing affects the incidence of aGVHD after allo-PBSCT. The primary endpoint will be the cumulative incidence of grade II to IV aGVHD within 100 days after transplantation. Acute GVHD will be graded according to the Mount Sinai Acute GVHD International Consortium criteria.

2) Secondary endpoints included the incidence of grade III to IV aGVHD within 100 days after transplantation, neutrophil engraftment, platelet recovery, chronic GVHD (cGVHD), TRM, overall survival (OS), disease-free survival (DFS), and GRFS (survival without III-IV aGVHD, cGVHD requiring systematic treatment, relapse or death).

3. **Study design**

3.1 Overall design

This is an investigator-initiated, multicenter, prospective, randomized, open-label, phase 3 clinical trial designed to evaluate the effect of stem cell infusion timing on the incidence and severity of aGVHD following allo-PBSCT, as well as its impact on long-term GRFS. Patients undergoing allo-PBSCT will be randomly assigned in a 1:1 ratio to receive stem cell infusion at either 12:00 pm (±30 minutes) or 6:00 pm (±30 minutes).

Table 1. Study Flow Diagram

| **Study period** | **Screening** | **Treatment** | **Follow-up** | | |
| --- | --- | --- | --- | --- | --- |
| **Visit** | **Screening** | **Stem cell infusion** | **Visit 1** | **Visit 2** | **Visit 3** |
| **Time window** | **Day -8 to -1 before transplantation** | **Day 0 (transplantation)** | **Day +30 (±3 days)** | **Day +100 (±7 days)** | **Day +360 (±7 days)** |
| **Informed consent** | **×** |  |  |  |  |
| **Demographics** | **×** |  |  |  |  |
| **Medical history** | **×** |  |  |  |  |
| **Concomitant medications** | **×** | **×** | **×** | **×** | **×** |
| **Vital Signs** | **×** | **×** |  |  |  |
| **Complete blood count** | **×** | **×** | **×** | **×** | **×** |
| **Blood biochemistry** | **×** | **×** | **×** | **×** | **×** |
| **Bone marrow examination** |  |  | **×** |  | **×** |
| **ECOG performance status** | **×** | **×** | **×** | **×** | **×** |
| **Eligibility assessment** | **×** |  |  |  |  |
| **Study intervension** |  | **×** |  |  |  |
| **GVHD assessment** |  |  | **×** | **×** | **×** |
| **Survival status** |  | **×** | **×** | **×** | **×** |

3.2 GVHD assessment criteria

3.2.1 Diagnostic and grading criteria for aGVHD

aGVHD will be diagnosed and graded according to the criteria established by the Mount Sinai Acute GVHD International Consortium (MAGIC). aGVHD typically occurs within 100 days after allogeneic hematopoietic stem cell transplantation and primarily involves the skin, gastrointestinal tract, and liver. Clinical manifestations include maculopapular rash, diarrhea, and cholestatic liver dysfunction. The diagnosis of upper gastrointestinal aGVHD requires confirmation by endoscopic evaluation, with or without histopathological findings.

**Table 2 MAGIC criteria for organ stage and overall grade in aGVHD**

| Stage | Skin (Active Erythema Only) | Liver (Total Bilirubin) | Upper GI | Lower GI |
| --- | --- | --- | --- | --- |
| 0 | None | <2 mg/dL | None or intermittent nausea/vomiting | Adults: <500 mL/day; <3/day  Children: <10 mL/kg/day or <4 episodes/day |
| 1 | <25% BSA | 2-3 mg/dL | Persistent nausea/vomiting | Adults: 500-999 mL/day; 3-4/day  Children: 10-19.9 mL/kg/day or 4-6 episodes/day |
| 2 | 25-50% BSA | 3.1-6 mg/dL | — | Adults: 1000-1500 mL/day; 5-7/day  Children: 20-30 mL/kg/day or 7-10 episodes/day |
| 3 | >50% BSA | 6.1-15 mg/dL | — | Adults: >1500 mL/day; >7/day  Children: >30 mL/kg/day or >10 episodes/day |
| 4 | Generalized with bullae | >15 mg/dL | — | Severe abdominal pain ± ileus/bloody stool |
| Grade |  |  |  |  |
| 0 | No organ involvement (all organs at stage 0) | | | |
| I | Stage 1-2 |  |  |  |
| Ⅱ | Stage 3 | Stage 1 | Stage 1 | Stage 1 |
| Ⅲ | Stage 0-3 | Stage 2-3 | Stage 0-1 | Stage 2-3 |
| Ⅳ | Stage 4 | Stage 4 | Stage 0-1 | Stage 4 |

Footnotes: For adults, 200 mL of stool output is considered equivalent to one episode; for children, 3 mL/kg is considered equivalent to one episode. The overall grade is determined based on the most severely affected target organ.

3.2.2 Diagnostic and grading criteria for cGVHD

cGVHD is diagnosed based on clinical manifestations rather than a strict time threshold, although it typically occurs beyond 100 days after transplantation. According to the National Institutes of Health (NIH) consensus criteria, the diagnosis of cGVHD requires the presence of at least one diagnostic manifestation or at least one distinctive manifestation confirmed by appropriate evaluation.

**Table 3 NIH 2014 criteria for cGVHD scoring and global severity**

| **Organ/System** | **Score 0** | **Score 1** | **Score 2** | **Score 3** |
| --- | --- | --- | --- | --- |
| **Performance Status** | Asymptomatic; fully active (ECOG 0) | Symptomatic; mild limitation (ECOG 1) | Symptomatic; ambulatory and capable of self-care, >50% out of bed (ECOG 2) | Symptomatic; limited self-care, >50% in bed (ECOG 3-4) |
| **Skin** |  |  |  |  |
| -% Body Surface Area (BSA) | No involvement | <18% BSA | 19–50% BSA | >50% BSA |
| -Skin Features | No sclerotic changes | — | Superficial sclerosis; mobile | Deep sclerosis; nonmovable, restricted mobility, or ulceration |
| **Mouth** | No symptoms | Mild symptoms; no limitation of oral intake | Moderate symptoms; mild limitation of oral intake | Severe symptoms; significant limitation of oral intake |
| **Eyes** | No symptoms | Mild dry eye; no impact on daily activities (≤3 lubricating drops/day) | Moderate dry eye affecting daily activities (>3 drops/day), without keratoconjunctivitis sicca (KCS) | Severe dry eye significantly affecting daily activities, or inability to work due to ocular symptoms, or vision impairment due to KCS |
| **Gastrointestinal Tract** | No symptoms | Symptoms without significant weight loss (≤5% within 3 months) | Mild–moderate weight loss (5–15% within 3 months) or moderate diarrhea without limitation of daily activities | Severe weight loss (>15% within 3 months), requiring nutritional support, or esophageal dilation, or severe diarrhea limiting daily activities |
| **Liver** | Normal TBIL; ALT or ALP <3 × ULN | Normal TBIL; ALT 3-5 × ULN or ALP ≥3 × ULN | TBIL ≤3 mg/dL (≤51.3 μmol/L) or ALT >5 × ULN | TBIL >3 mg/dL (>51.3 μmol/L) |
| **Lung** |  |  |  |  |
| - Symptoms | None | Mild (dyspnea when climbing 1 flight of stairs) | Moderate (dyspnea on flat ground) | Severe (dyspnea at rest or requiring oxygen) |
| -%FEV1 | ≥80% | 60-79% | 40–59% | ≤39% |
| **Joints and Fascia** | No symptoms | Mild tightness; normal or mildly reduced ROM; no impact on daily activities | Tightness or contracture; fasciitis-related erythema; moderate ROM reduction; mild-moderate impact | Contracture with severe ROM limitation; major impairment in daily activities (e.g., unable to tie shoes or button clothing) |
| **Genital Tract** | No symptoms | Mild symptoms; no significant discomfort on examination | Moderate symptoms; mild discomfort on examination | Severe symptoms |
| **Global Severity of cGVHD** | No evidence of cGVHD | Mild  1-2 organs involved with score 1; lung score must be 0 | Moderate  ≥3 organs with score 1, or ≥1 organ (excluding lung) with score 2, or lung score = 1 | Severe  ≥1 organ (excluding lung) with score 3, or lung score ≥2 |

3.3 Assessment of Transplantation-Related Complications

Transplantation-related complications and adverse events will be systematically monitored throughout the study. Adverse events will be recorded and graded according to the National Cancer Institute Common Terminology Criteria for Adverse Events (CTCAE), version 5.0. Safety assessments will focus on transplantation-related complications and infectious events. No intervention-specific toxicities are expected, as the study intervention involves only the timing of stem cell infusion.

4 **Patient eligibility**

**4.1 Inclusion criteria**

1) Definite diagnosis of malignant hematologic disease before transplantation,

2) age 12-60 years old, gender is not limited, race is not limited;

3) Patients who are proposed to receive allo-PBSCT for the first time;

4) Eastern Cooperative Oncology Group (ECOG) score 0-2;

5) No serious organ failure or active infection;

6) Voluntary open randomized controlled study to observe whether the time of stem cell infusion affects the occurrence of aGVHD after transplantation;

7) Each subject must sign an informed consent form (ICF) indicating that he/she understands the purpose and procedures of the study and is willing to participate in the study; in view of the patient's condition, if the patient's signature is not conducive to the treatment of his/her condition, the ICF will be signed by the legal representative.

**4.2 Exclusion criteria**

1) Severe organ dysfunction or disease, such as severe disease and dysfunction of the heart, liver, kidneys and pancreas;

2) Pregnant patients;

3) Patients and/or authorized family members who refuse to undergo an open randomized controlled study to observe whether the time of stem cell infusion affects the occurrence of aGVHD after transplantation;

4) Any life-threatening disease, physical condition, or organ system dysfunction that, in the opinion of the investigator, may compromise patient safety and put the results of the study at unnecessary risk; drug-dependent individuals; patients with uncontrolled psychiatric disorders; and individuals with cognitive dysfunction;

5) Participants in other clinical studies that may affect aGVHD within 3 months;

6) Those whom the investigator considers unsuitable for enrollment (e.g., those who anticipate that patients will not be able to adhere to the examination and treatment due to financial and other issues).

**4.3 Withdrawal and discontinuation criteria**

Participants may be withdrawn from the study under the following circumstances:

1) Withdrawal of informed consent by the participant or their legal representative;

2) Loss to follow-up or inability to continue study assessments;

3) Occurrence of clinical conditions unrelated to the study intervention that preclude continued participation;

4) Death from any cause;

5) Any other reason that, in the judgment of the investigator, makes continued participation inappropriate.

**5. Intervention**

After enrollment, participants will be randomly assigned to either the early stem cell infusion group or the late stem cell infusion group. Participants in the early infusion group will receive stem cell infusion at 12:00 pm (±0.5 hour), whereas those in the late infusion group will receive stem cell infusion at 6:00 pm (±0.5 hour) on transplantation day 0. All other procedures will be performed according to standard allo-PBSCT protocols at each participating center. Following collection, the graft will be evaluated for CD34⁺ cell content by flow cytometry. If the infused CD34⁺ cell dose is less than 4 × 10⁶ cells/kg of recipient body weight, an additional stem cell infusion may be administered on the following day within the same assigned time window.

6. **Research process**

6.1 Recruitment and baseline assessment

This is a multicenter, prospective, randomized phase 3 clinical trial designed to evaluate the effect of stem cell infusion timing on clinically relevant outcomes following allo-PBSCT. The indication for allogeneic transplantation will be determined by the treating physician according to institutional standards. Patients will be evaluated at each participating center and counseled regarding the risks and benefits of allo-PBSCT as part of routine clinical care. Participants who meet the eligibility criteria specified in **Section 4.1** will undergo baseline evaluations prior to transplantation. These assessments will serve as reference measures for subsequent clinical analyses. Baseline assessments will include, but are not limited to, the following:

1) Physical examination (including height, weight, and body surface area)

2) Vital signs (temperature, heart rate, and blood pressure)

3) Complete blood count (CBC)

4) Blood biochemistry (including liver and renal function)

5) Coagulation profile

6) Bone marrow examination (aspirate and/or biopsy, as clinically indicated)

7) Minimal residual disease (MRD) assessment and relevant molecular testing

8) Imaging studies (e.g., ultrasound, CT, or MRI), as clinically indicated

6.2 Registration and informed consent

Upon confirmation of eligibility, participants will be formally enrolled and assigned a unique study identification number through the online study data management system. The principal investigator and study team will be notified upon completion of enrollment. All participants must provide written informed consent before initiation of any study-specific procedures. Participants will be fully informed about the nature of their disease, the study objectives, procedures, potential risks and benefits, alternative treatment options, and the voluntary nature of participation. Adequate time will be provided for questions and consideration prior to consent. The informed consent process will be conducted in accordance with institutional review board-approved procedures and applicable regulatory requirements. The following documents will be obtained prior to study enrollment: 1) completion of screening assessments; 2) signed written informed consent; and 3) documentation confirming eligibility for allo-PBSCT.

6.3 Transplantation procedures

6.3.1 Conditioning regimens

Conditioning intensity (myeloablative conditioning [MAC] or reduced-intensity conditioning [RIC]) will be selected at the discretion of the treating physician, taking into account patient- and disease-related factors. Risk stratification may be guided by established tools such as the Hematopoietic Cell Transplantation-Comorbidity Index (HCT-CI) and the Disease Risk Index (DRI). Pretransplant evaluation will include bone marrow examination performed within 28 days prior to transplantation (day 0), along with standard clinical assessments of organ function, including cardiac, pulmonary, hepatic, and renal evaluation, according to institutional practice. Minimal residual disease (MRD) assessment, molecular testing, and chimerism analysis may be performed at the discretion of the treating center and are not mandated by the study protocol. Common regimens consist of busulfan-based combinations with cyclophosphamide or melphalan, often incorporating fludarabine or cytarabine, as well as total body irradiation (TBI) or total marrow irradiation (TMI)-based regimens with cyclophosphamide and/or additional cytarabine. Specific drug dosing and schedules will follow institutional standards and are not dictated by the study protocol. Specific drug dosing schedules, administration timing, and supportive care measures will follow institutional standards. Minor adjustments to the conditioning schedule are permitted to accommodate clinical or logistical considerations.

Commonly used MAC regimens include the following:

1) Busulfan combined with cyclophosphamide (Bu/Cy): intravenous busulfan 3.2 mg/kg/day for 3 - 4 days and cyclophosphamide 60 mg/kg/day or 1.8 g/m^2^/d for 2 days supplemented with fludarabine (Flu) (30 mg/m^2^ daily for 4 - 5 days) or aytarabine (Ara-C) added (2 g/m^2^ daily for 4 days);
2) Busulfan combined with melphalan (Bu/Mel): intravenous busulfan 3.2 mg/kg/day for 3 - 4 days and melphalan 50-60 mg/kg/day for 2 days supplemented with fludarabine (Flu) (30 mg/m^2^ daily for 4 - 5 days) or aytarabine (Ara-C) added (2 g/m^2^ daily for 4 days);
3) Total body irradiation (TBI) or total marrow irradiation (TMI)-based regimens in combination with cyclophosphamide: fractionated TBI (total 12 Gy, 4 fractions) or TMI (total 15 Gy, 3 fractions) and Cy, 40 - 60 mg/kg daily for 2 days).

Commonly used RIC regimens include the following:

1) Busulfan combined with melphalan (Bu/Mel): intravenous busulfan 3.2 mg/kg/day for 2 days and melphalan 50 mg/kg/day for 2 days supplemented with fludarabine (Flu) (30 mg/m^2^ daily for 5 days).

Specific drug dosing schedules, administration timing, and supportive care measures will follow institutional standards. Minor adjustments to the conditioning schedule are permitted to accommodate clinical or logistical considerations.

6.3.2 GVHD prophylaxis regimens

All patients will receive calcineurin inhibitor (CNI)-based prophylaxis combined with mycophenolate mofetil (MMF) as the backbone regimen. In addition to this standard platform, prophylaxis may be intensified according to donor type and institutional practice with one of the following approaches: 1) rabbit anti-thymocyte globulin (rATG) or anti-T-lymphocyte globulin (ATLG); 2) posttransplant cyclophosphamide (PT-Cy); 3) combination of ATG and PT-Cy; and 4) optional addition of short-course methotrexate (MTX). The choice of regimen will be determined by the treating center and is not restricted by the study protocol. No study-specific interventions beyond the randomized timing of stem cell infusion will be needed.

6.3.2.1. Cyclosporine

Cyclosporine will be initiated on day -1 at a starting dose of 2-2.5 mg/kg/day administered as a continuous intravenous infusion and subsequently adjusted to achieve target trough levels of 250-350 ng/mL. Serum cyclosporine levels will be monitored at least twice weekly during hospitalization and thereafter according to institutional practice. Cyclosporine may be transitioned to oral administration once the patient is able to tolerate oral intake. Concurrent use of azole antifungal agents (e.g., itraconazole, voriconazole, or fluconazole at doses >200 mg/day) may inhibit cyclosporine metabolism and increase drug levels. Dose adjustments and more frequent monitoring are recommended when these agents are coadministered.

6.3.2.2. Tacrolimus

Tacrolimus will be initiated on day -3 at a starting dose of 0.03 mg/kg/day administered as a continuous intravenous infusion and adjusted to maintain target trough levels of 5-10 ng/mL. Drug levels will be monitored at least twice weekly during hospitalization and thereafter according to institutional practice. Tacrolimus may be transitioned to oral administration once tolerated. Azole antifungal agents may increase tacrolimus exposure; therefore, dose reduction and enhanced monitoring are recommended when used concomitantly.

6.3.2.3. MMF

MMF will be initiated on day +1 at a dose of 1.0 g orally twice daily or 25-30 mg/kg/day divided into two doses. MMF will typically be tapered beginning around day +21 and discontinued by approximately 2 months post-transplant, according to institutional practice.

6.3.2.4. rATG or ATLG

rATG may be administered as part of the conditioning or prophylaxis regimen at a total dose of 4.5-7.5 mg/kg given from day -4 to day -2. In selected cases, posttransplant administration of rATG with a total dose of 2.5 mg/kg on day +15 or +16 may be administered according to institutional protocols. ATLG may be administered at a total dose of 10-25 mg/kg from day -4 to day -2, according to institutional practice.

6.3.2.5. PT-Cy

PT-Cy will be administered on days +3 and +4 at a total dose of 60-100 mg/kg, in accordance with institutional protocols. Intravenous mesna will be administered for uroprotection in all patients receiving cyclophosphamide, including both conditioning and PT-Cy phases.

6.3.2.6. MTX

Methotrexate may be administered as short-course prophylaxis at 15 mg/m² on day +1 and 10 mg/m² on days +3 and +6. The day +1 dose should be administered at least 24 hours after stem cell infusion. Folinic acid rescue may be administered on days +1, +3, and +6, beginning approximately 12 hours after methotrexate infusion, according to institutional practice.

6.3.3 Donor preparation and stem cell collection

Donors will be selected based on standard high-resolution HLA typing, including HLA-A, -B, -C, -DRB1, and -DQB1 loci. Donor eligibility assessment and counseling regarding the risks of granulocyte colony-stimulating factor (G-CSF) administration and apheresis will be conducted according to institutional guidelines. Written informed consent will be obtained from all donors prior to stem cell collection. Peripheral blood stem cells will be mobilized using G-CSF and collected via apheresis according to standard institutional procedures. The target CD34⁺ cell dose for transplantation will typically range from 2 × 10⁶ to 10 × 10⁶ cells/kg of recipient body weight. Excess collected cells may be cryopreserved for potential donor lymphocyte infusion (DLI) according to institutional practice.

6.3.4 Treatment monitoring protocols

1) Monitoring indicators before hematopoietic reconstruction:

(1) Changes in body temperature, pulse, and blood pressure (daily)

(2) Routine blood examination (twice a week)

(3) Liver, kidney, and coagulation function tests (weekly)

(4) CMV, EB and other viral indicators (1-2 times a week)

2) After hematopoietic reconstitution to 1 year of transplantation:

(1) Routine blood test, liver and kidney function test, CMV and EB and other viral indicators (at least monthly)

(2) Bone marrow evaluation (aspirate and/or biopsy): as clinically indicated

(3) Minimal residual disease (MRD) assessment and molecular testing: periodically according to institutional practice.

7. **Study endpoints**
   7.1 Primary study endpoints

1) The cumulative incidence of grade II-IV aGVHD at 100 days post-transplantation. Acute GVHD was graded according to the MAGIC criteria.

7.2 Secondary study endpoints

1) The cumulative incidence of grade III to IV aGVHD in the first 100 days post-transplant. aGVHD was graded according to the Mount Sinai Acute GVHD International Consortium criteria.

2) The cumulative incidence of neutrophil engraftment at 28 days after transplantation. Neutrophil engraftment time was defined as the first of three consecutive days during which the neutrophil count was at least 0.5×10^9/L.

3) The cumulative incidence of platelet recovery at 100 days after transplantation. Platelet engraftment is defined as independence from platelet transfusion for at least 7 days with a platelet count of 50×10^9/L.

4) The cumulative incidence of transplant-related mortality at 180 days after transplantation.

5) The cumulative incidence of transplant-related mortality at 360 days after transplantation.

6) The cumulative incidence of cGVHD at 360 days after transplantation. The severity of chronic GVHD was graded according to the 2014 NIH criteria.

7) The probability of GRFS at 360 days after transplantation. The composite endpoint of GRFS was defined as the first events occurring after transplantation among grade III to IV aGVHD, moderate to severe cGVHD, relapse, or death for any reason.

8) The probability of disease-free survival (DFS) at 360 days after transplantation.

9) The probability of OS at 360 days after transplantation.

8. **Statistical analysis plan**
   8.1 Sample size calculation

Sample size estimation was based on the primary end point, defined as the incidence of grade II to IV aGVHD within 100 days after transplantation. Based on our preliminary single-center retrospective cohort of 134 patients undergoing matched sibling donor transplantation, in which the incidence of grade II to IV aGVHD was 18.0% among patients receiving stem cell infusion before 2:00 pm and 41.0% among those receiving infusion at or after 2:00 pm. Assuming a two-sided α level of 0.05 and a statistical power of 90%, a total sample size of 178 patients (89 per group) was required to detect a between-group difference. To account for potential attrition due to early death, relapse or other unforeseen events, the sample size was increased by 10%, resulting in a planned enrollment of 198 patients. Sample size calculations were performed using G*Power software.

8.2 Randomization and statistical analysis

Randomization was performed centrally by independent statisticians using a computer-generated sequence with randomly varying permuted block sizes of 4, 6, and 8, stratified according to the study center. Eligible participants who provided written informed consent were assigned in a 1:1 ratio to the 12:00 pm infusion group or the 6:00 pm infusion group. Allocation was concealed and implemented through an electronic data capture system. Statisticians involved in randomization were not involved in patient recruitment, treatment, or outcome assessment.

The primary end point, defined as the cumulative incidence of grade II to IV aGVHD within 100 days after transplantation, was analyzed using a competing risk framework, with death without prior aGVHD considered a competing event. Cumulative incidence functions were estimated and compared using Gray’s test, and subdistribution hazard ratios (HRs) with 95% confidence intervals (CIs) were calculated using the Fine-Gray subdistribution hazard model. Prespecified multivariable analyses will be conducted to adjust for potential confounding factors. Covariates will be selected based on clinical relevance and may include baseline demographic and transplantation-related characteristics including age, sex, disease diagnosis, disease status at transplantation, conditioning intensity (myeloablative vs reduced-intensity conditioning), graft types, infused CD34^+^ cell dose, infused TNCs dose, donor-recipient HLA compatibility, donor-recipient sex compatibility (female donor to male recipient vs others), GVHD prophylaxis regimen, and methotrexate use. Additional variables may be included as appropriate based on data availability and clinical considerations.

Secondary end points, including grade III to IV aGVHD, cGVHD, relapse, and TRM, were analyzed using similar competing risk approaches, with appropriate competing events specified for each outcome. Time-to-event outcomes without competing risks, including OS, DFS, and GRFS, were estimated using the Kaplan‒Meier method and compared between groups using the log-rank test.

All statistical tests were two-sided, with a significance level of 0.05. Statistical analyses were performed using R software (version 4.2.2, R Foundation for Statistical Computing, Vienna, Austria).

8.3 Analysis populations

Full Analysis Set (FAS)

The full analysis set will be defined according to the intention-to-treat (ITT) principle and will include all randomized participants. Participants will be analyzed according to their assigned group, regardless of protocol adherence or completion of treatment. The FAS will serve as the primary population for efficacy analyses.

Per-protocol set (PPS)

The per-protocol set will include participants who adhere sufficiently to the study protocol without major protocol deviations. Major protocol deviations will be prespecified and may include failure to meet key eligibility criteria, substantial noncompliance with study procedures, or deviation from the assigned infusion time window. The PPS will be used for sensitivity analyses to assess the robustness of the primary findings.

9. **Ethics and regulatory considerations**

This clinical trial will be conducted in accordance with the Declaration of Helsinki, the International Council for Harmonization Good Clinical Practice (ICH-GCP) guidelines, and all applicable local regulatory requirements.

9.1 Ethics committee approval

The study protocol, informed consent forms, and all relevant study documents will be reviewed and approved by the institutional review board or ethics committee at each participating center prior to study initiation. Any protocol amendments, including changes to the informed consent form, will require prior approval by the ethics committee before implementation.

9.2 Informed consent

Written informed consent will be obtained from all participants or their legal representatives prior to the initiation of any study-specific procedures. Participants will be provided with sufficient information regarding the study objectives, procedures, potential risks and benefits, and alternative treatment options to enable an informed decision. The informed consent process will be conducted by qualified study personnel in accordance with IRB-approved procedures.

9.3 Data sharing

Individual participant data may be made available upon reasonable request after completion of the study and publication of the primary results, in accordance with institutional and regulatory policies.

**10. Study management**

Participants will be monitored throughout the study by trained investigators. Patients will have access to study personnel for reporting adverse events or concerns at any time. Participant adherence will include compliance with scheduled visits, study procedures, and follow-up assessments. Participants may be withdrawn from the study in cases of poor adherence, withdrawal of consent, or at the discretion of the investigator for safety or other clinical reasons.

**11. Data management**

11.1 Confidentiality

All participant information will be kept confidential in accordance with applicable data protection regulations. Participants will be identified by unique study identification numbers, and no personally identifiable information will be disclosed in study reports. Access to study data will be restricted to authorized personnel, including investigators and regulatory authorities, for purposes of monitoring, auditing, and inspection.

11.2 Record retention

All study-related documents, including source data, case report forms (CRFs), informed consent forms, and regulatory documents, will be maintained in accordance with applicable regulatory requirements and institutional policies. Records will be retained for a minimum period required by national regulations and institutional guidelines.

11.3 Protocol amendments

The study protocol will not be modified after approval except under exceptional circumstances. Any protocol amendments will be documented, justified, and submitted for approval to the ethics committee prior to implementation. Amendments may be considered in the following situations:

1) Difficulty in participant recruitment requiring modification of eligibility criteria;

2) New evidence or interim findings indicating the need to revise study assumptions or sample size.

All amendments will require approval by the principal investigator and ethics committee.

**12. Quality control and assurance**

12.1 Compliance with protocol and GCP

The study will be conducted in accordance with the approved protocol and GCP guidelines. All study personnel will be trained in protocol procedures prior to study initiation.

12.2 Monitoring

The study will be monitored to ensure compliance with the protocol, GCP, and regulatory requirements. Monitoring activities will include verification of informed consent, source data review, and assessment of protocol adherence.

12.3 Data quality assurance

All study data will be collected, managed, and analyzed using standardized procedures to ensure accuracy, completeness, and reliability. Data verification procedures will be implemented to ensure that the reported results are consistent with the source data.

12.4 Audits and Inspections

The study may be subject to audit by the sponsor or inspection by regulatory authorities. Investigators agree to provide direct access to study documents and facilitate such audits or inspections in accordance with applicable regulations.
